# Hypermobility and Injury Among Instrumental Musicians: A Scoping Review

**DOI:** 10.1101/2025.08.13.25333390

**Authors:** Michelle L. King, Heather M. Macdonald

**Affiliations:** Radford University-Adjunct, (435)227-5081; Oklahoma State University-Visiting

## Abstract

**Objective:** Up to 86% of musicians experience playing-related musculoskeletal problems (PRMPs). Joint hypermobility (JH), which affects up to 34% of the population, may be a risk factor for such injuries, however, research on this topic is limited. The aims in this scoping review are to: (1) map information in existing literature on the relationship between JH and PRMPs in instrumental musicians, (2) identify subpopulations at risk of JH-related injuries, and (3) map supportive strategies used to accommodate hypermobile instrumentalists.

**Design:** The review was conducted following JBI methodology and adapted for a master’s thesis. Searches were performed in MEDLINE, Music Index, SPORTDiscus, and gray literature databases, using keywords related to “instrumental musicians” and “hypermobility,” resulting in 1570 sources.

**Results:** Of 165 relevant sources, 79 included original data on hypermobility, with only 30 primarily focused on JH. Most sources were published among populations primarily of European descent and adults ages 18-40 in professional or post-secondary classical settings. Research gaps identified include studies addressing hormonal influences on joint laxity, non-European populations, children, amateur musicians, and neurodivergent individuals. Sources containing original JH information consisted of 45% empirical studies (mostly prevalence) and 55% anecdotal reports. In 72% of all sources, authors concluded JH negatively impacts musicians.

**Conclusions:** Inconsistent results among empirical studies and incongruences between results and anecdotal evidence are indicative of methodological weaknesses. Limitations in measurement tools were noted, affecting study design and data interpretation. Future researchers should conduct qualitative research to capture experiences of hypermobile musicians to inform study design. They should expand quantitative methods, particularly longitudinal and randomized controlled trials, and incorporate sensitive, joint-specific assessments. Training for healthcare professionals, musicians, and music teachers should include JH and health impacts on musicians to ensure accurate research design and interpretation.

**Registration:** The scoping review protocol (https://osf.io/c5rzn) was previously registered and published on the Open Science Framework (OSF) along with all official documents, search query strings, and raw data (https://osf.io/6jynk/).

**STRENTHS AND LIMITATIONS OF THIS STUDY:**

- This is the first review focused on instrumental musicians with joint hypermobility.
- A thorough and detailed search strategy was developed with the assistance of expert research librarians and the review was conducted in accordance with the JBI scoping review manual and PRISMA-Scr guidelines, except where noted.
- The review was conducted as a master’s thesis requirement for Radford University. Due to time and resource constraints, only one reviewer was involved in data extraction and analysis with oversight of a research committee.
- As is standard with scoping reviews, sources were not evaluated for quality, therefore, conclusions cannot be generalized.

## Background

According to a survey performed by the National Association of Music Merchants, in 54% of American households at least one person plays a musical instrument^1^. However, up to 86% of people who play a musical instrument experience playing-related pain and injury.^2^ Playing-related musculoskeletal problems (PRMPs), defined as any” pain, weakness, numbness, tingling, or other symptoms that interfere with [the] ability to play [an] instrument at the level [one is] accustomed to,”^3^[p293)] are problematic because they develop over time, and by the time a person seeks treatment, the consequences are often extreme or irreversible.^4^ PRMPs have implications beyond inability to perform music, causing loss of employment, income, opportunity, and career progression.^4^ The injury may cause pain and difficulty in completing daily living tasks such as driving, cleaning, or caring for children.^5^ PRMPs have also been shown to have a negative effect on musicians’ mental health.^6^

One potential risk factor for PRMPs is the presence of joint hypermobility (JH), which is found in up to 34.1% of the general population.^7,8^ Among the general population, injury prevention efforts directed toward treating JH are effective in reducing the prevalence of injuries.^9^ Although the research regarding musicians with JH is sparse and inconclusive,^8^ it is suggestive that addressing JH among musicians could result in similar injury reduction.^10^

Medical practitioners have a responsibility to understand unique risks and use appropriate practices when treating musicians with JH. However, in a preliminary review of peer reviewed literature, very few sources and no randomized controlled trials or intervention studies related to instrumental musicians with JH were found. This lack of research limits medical practitioners’ ability to use evidence-based approaches when treating musicians with JH. The aim of this study was to identify existing gaps in research through an extensive review to guide appropriate design for future studies, and answer the following research questions:

1. What is known from the literature about the relationship between hypermobility and playing-related injury and difficulty among instrumental musicians?
2. Are there subpopulations of instrumental musicians with an elevated risk of playing-related injury and difficulty due to hypermobility?
3. What supportive methods have been used for instrumental musicians with hypermobility?

## Methods

This review was conducted through the perspective of Pragmatism with the purpose of evaluating knowledge in terms of function and consequences for musicians with JH.^11^ It was conducted in accordance with the JBI methodology for scoping reviews^12^ using the Preferred Items for Systematic Reviews and Meta-analysis Protocols.^13^ The required PRISMA-ScR Checklist is included (online supplemental appendix A; figure A1). The scoping review protocol^14^ was previously registered and published on the Open Science Framework (OSF) along with all official documents, search query strings, and raw data collected for the review.^15^

### Data sources and search strategy

#### Preliminary search

In accordance with JBI protocol, a preliminary search of MEDLINE, Music Index, PROSPERO International Prospective Register for Systematic Reviews, Cochrane Database of Systematic Reviews, JBI Systematic Review Register, and JBI Evidence Synthesis was conducted. No systematic reviews, nor scoping reviews on JH among musicians were identified. Next, a limited search of Music Index was undertaken to identify articles on musicians with JH. Keywords from titles and abstracts of relevant articles, and index terms listed in abstracts were used to develop a full search strategy.

#### Official search

The search strategy included all previously identified keywords and index terms. Because a scoping review is iterative^12^, and the Medical Subject Headings (MeSH) are unique to each database, the search strategy was adapted for each database. A search of the International Clinical Trials Registry Platform (ICTRP),^16^ clinicaltrials.gov^17^ were also searched to track publication bias by comparing registered trials to published trials. The reference lists of all included sources were screened for additional studies. Additional sources the author encountered that met inclusion criteria were also added.

The research question involves both medical and music topics, thus MEDLINE and Music Index with Full Text, which are the most encompassing and recommended databases in their respective fields^18,19^ were searched. Because performing arts medicine also falls under the category of sports medicine, SPORTDiscus, the leading database for sports medicine,^20^ was searched. For the complete search strategy see online supplemental appendix B, Table B1.

### Title and abstract screening

#### Inclusion and exclusion criteria

A scoping review should be as comprehensive as possible,^12^ therefore, all types of available sources, regardless of research type, peer-review, date of authorship, or publication status, including opinion papers, were included. Sources in which any population such as age, biological sex, socioeconomic status, and health condition were included as well as those from all instrumental contexts and locations. This included but was not limited to educational and professional ensemble groups, beginning and amateur musicians, and both western and non-western musicians. Only articles in English were included.

To answer the primary review question, “What is known from the literature about the relationship between hypermobility and playing-related injury and difficulty among instrumental musicians,” only articles in which both musicians and Joint Hypermobility (JH) are discussed were included. To answer the question, “Are there subpopulations of instrumental musicians with an elevated risk of playing-related injury and difficulty due to hypermobility,” information related to subgroups such as females, youth, and neurodivergent people were collected. To answer the question, “What supportive methods have been used for instrumental musicians with hypermobility,” supportive techniques for people with JH were recorded.

Articles that did not mention both hypermobility and instrumental musicians were excluded. Although vocalists with JH may need specific support,^21^ technique differences between instrumental and vocal musicians warrant isolated research for each. To maintain a clear focus, articles in which only vocal musicians or non-musicians are discussed were not considered. Articles not in English were excluded to avoid translation errors and misunderstandings.

Initially, the JBI System for the Unified Management, Assessment and Review of Information (SUMARI)^22^ was proposed for managing review sources. However, custom forms could not be created within SUMARI; therefore, Covidence Systematic Review Software was selected as a replacement.^23^ Screening results were entered into a PRISMA Flow diagram. To comply with recommendations on the Open Science Framework platform and ensure consistency among reviewers, documents containing instructions for screening were created for and registered with the protocol. These documents can be accessed at https://osf.io/6jynk/files/osfstorage > Screening Documents.^15^

A total of 1,672 sources, which can be accessed at https://osf.io/6jynk/files/osfstorage > Official Search Results,^15^ were imported into Covidence following the database search. An additional 113 sources were identified through citation and gray literature searches, bringing the total to 1,785 sources. Two hundred and fifteen duplicates were removed, leaving 1,570 sources for title and abstract screening, completed by MK and HM. One thousand nine sources were excluded at this stage, and the full text of each of the remaining 561 sources was sought for retrieval. Fourteen sources could not be retrieved. Full texts of the remaining 547 sources were assessed for eligibility.

#### Full text screening

MK primarily conducted full text screening with some assistance from HM, MM, AD, and JB; not all sources were screened by two reviewers and final decisions were made by MK. Three hundred eighty-two sources were excluded for the following reasons: 370 were unrelated to JH; four were not in English; three were unrelated to either JH or instrumental musicians; two were not the source indicated; two contained discussion on JH and instrumental musicians separately, but not in conjunction; and one was not about instrumental musicians. Excluded sources and reasons for exclusion can be accessed at https://osf.io/56nx2.^15^ Ultimately, 165 sources were included in the review (figure 1). Documents containing instructions for screening were created for and registered with the protocol and can be accessed at https://osf.io/6jynk/files/osfstorage > Extraction Documents.^15^

**Figure 1.**
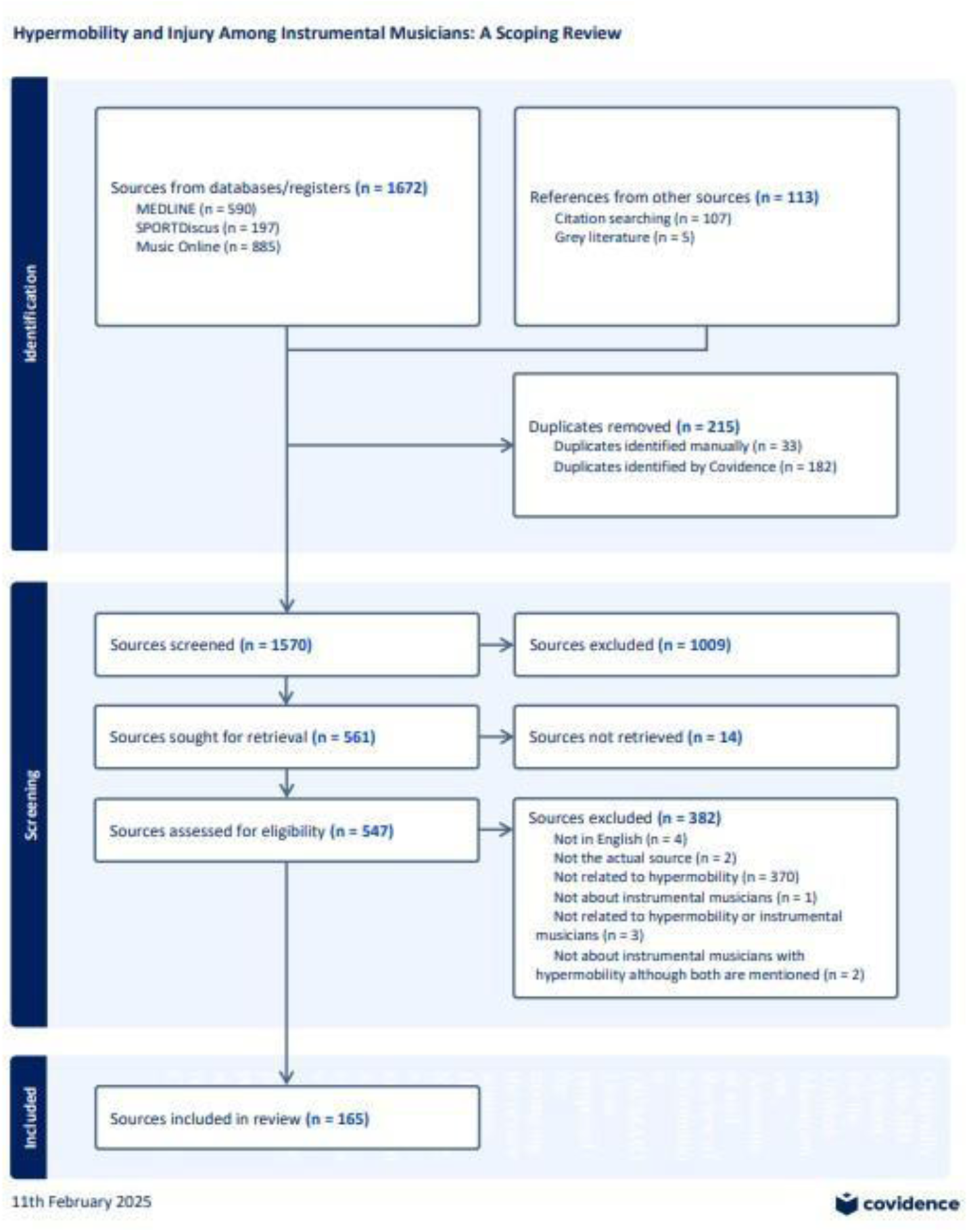
PRISMA flow chart

#### Data extraction

Data were extracted using a tool adapted from the JBI Extraction Form^12^ (online supplemental appendix C; table C1). In accordance with JBI recommendations, the draft extraction tool from the protocol was modified to enhance analytical precision and align with the extraction model in Covidence (online supplemental appendix C; figure C2).

Additional categories were incorporated to capture detailed source characteristics, including publication type and study design, facilitating a more accurate assessment of research gaps. Sections addressing recommendations for physicians and teachers, health and functional playing-related effects, prevalence data, factors influencing laxity, and research limitations were added to accommodate the extensive information available on these topics. Documents containing instructions for extraction were created for and registered with the protocol. These documents can be accessed at https://osf.io/6jynk/files/osfstorage<u>#</u> > Extraction Documents.^15^

#### Analysis and presentation of evidence

Although critical appraisal of individual evidence sources is not a requirement for scoping reviews,^24^ the registered protocol included plans to conduct critical appraisal, but was not completed in full or included in the analysis or report due to several limitations. First, the JBI tools^25^ used are designed for peer-reviewed sources and are not applicable to many review sources. Second, additional training in methodological procedures and data analysis would have been necessary to complete accurate analysis. Third, the scoring system created for the protocol involved assigning numerical values to the JBI tools, but it was not validated and did not accurately reflect the quality of the sources. Although not included in the analysis, appraisal scores and conversion tools may be accessed at https://osf.io/6jynk/files/osfstorage > Critical Appraisal Scores.^15^

#### Data charting process

Information was charted using two formats. The first involved aggregate data from all sources or from targeted lists derived from categories established by analyzing the extent to which JH was discussed in the literature and whether the information was original or cited and was analyzed using descriptive statistics. This data includes source relevance, origin, study type, aims, populations, and settings. The second format involved extracting specific findings and recommendations within each source. While some aggregate data is included, this format is focused on specific details to enhance understanding of the evidence. These findings include the relationship between JH and PRMPs, and reported impacts of JH on musicians. Information was also collected on supportive techniques, and research limitations identified by source authors.

## Results

Sources were categorized by the extent to which JH was discussed and the originality of data. These data are presented in Figure 2. First, source relevance was considered. Consistent with the preliminary search, information mentioning instrumental musicians with JH was scarce, comprising about 10% of available literature (*N* = 1570).

**Figure 2.**
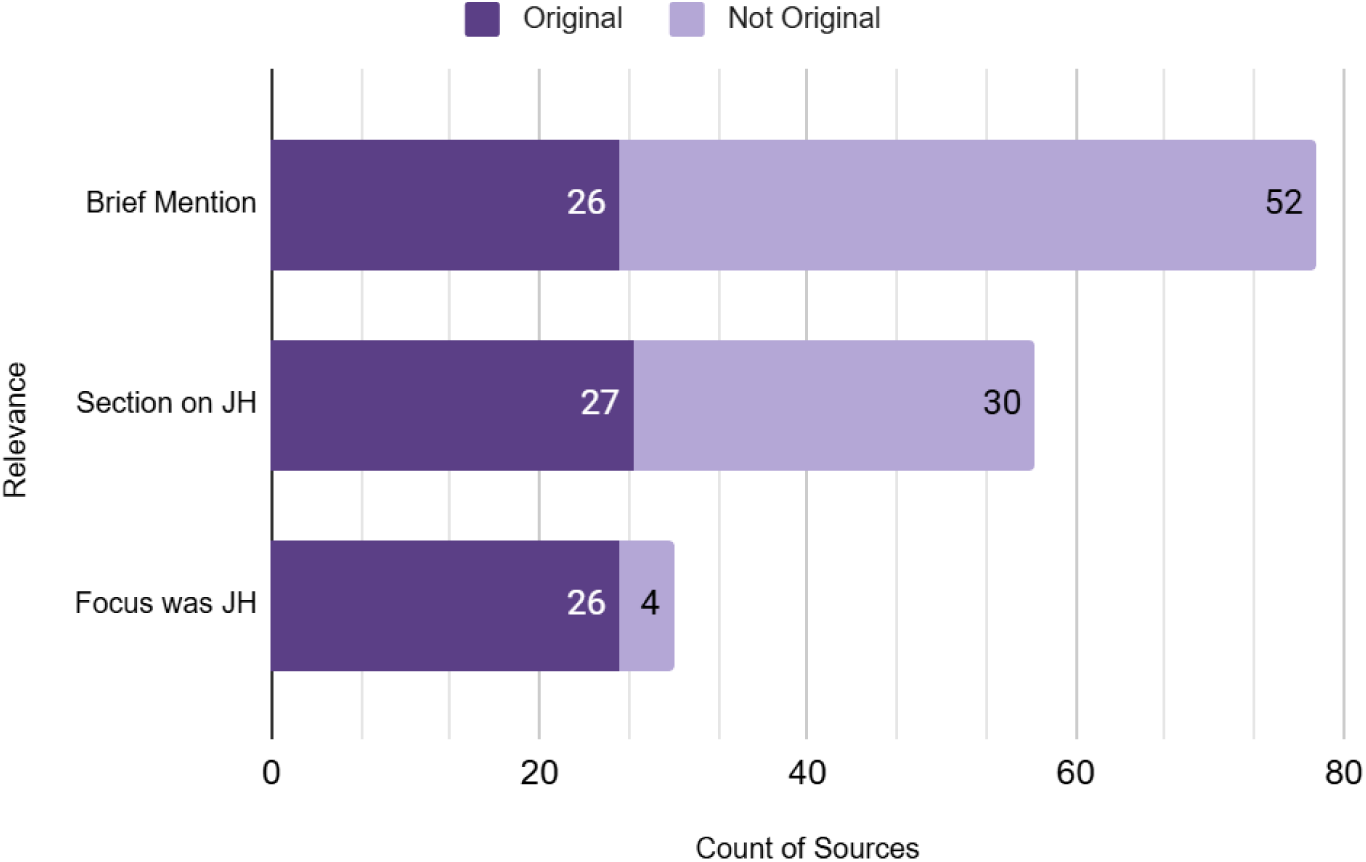
JH relevance and originality of sources

Next, source originality was assessed (figure 3) and three lists were derived from this information. For a complete list of sources in each list see online supplemental appendix D; tables D1-D3):

- **L1:** The primary list which includes all review sources–those in which both JH and instrumental musicians were mentioned (*N* = 165; 10.5%)
- **L2:** sources containing original information on JH (*n* = 79; 5.0%), compiled to prevent analysis of duplicated data
- **L3:** sources focused exclusively on JH (*n* = 30; 1.9%)

**Figure 3.**
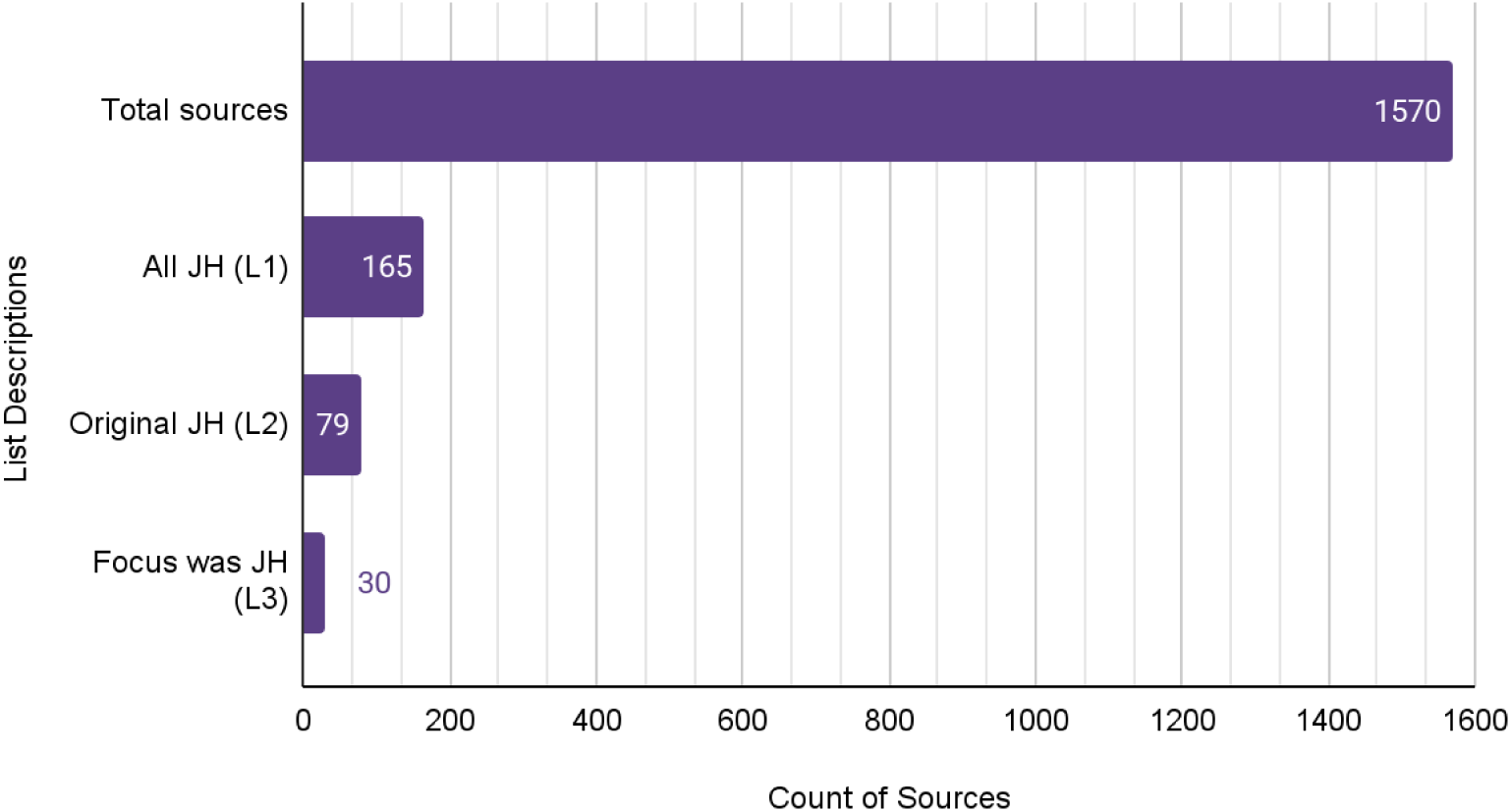
Derived lists

### Source characteristics

The publication date of the all sources ranged between 1947 and 2024, with most being published after 1980. In all three lists, most sources originated from countries in which the majority of individuals are of European descent such as the United States (US),^10,26–101^ Canada,^3,102–105^ the United Kingdom (UK),^106–131^ and Europe.^107,117,127,132–161^ Other locations of origin included Australia,^8,106,162–172^ Mediterranean,^173–175^ Russia,^176^ South Africa,^109,177,178^ China,^179^ India,^180^ and Brazil^181^. Some sources, such as systematic reviews or sources using online queries, were not reliant on a specific location or did not mention one.^182–184^ Mediterranean countries were grouped separately from the rest of Europe due to the higher reported prevalence of JH among those populations (online supplemental appendix E; table E1).

To avoid duplication of data, L1 was not counted for age distributions, performance settings, music genres, or instruments performed. Because joint laxity is influenced by age, the age distribution of subjects in L2 and L3 were charted to examine age-related trends in JH data and priorities among researchers. Most sources included adults ages 18-40. ^3,8,28,38,43,46,52,57,61–63,71,77,79,80,84,98,100–102,107–110,113,122,128,132,136,142,144,151,165,166,171,172, 184–186^ About one-third included adults over age 40.^28,46,57,61–63,79,102,108–110,128,136,165,171,184^ Less than one-third included children under age 18.^8,41,43,46,50,57,61,62,68,93,99,165,166,172,185^ Nearly half of sources did not specify an age ^26,27,27,29,35–37, 39,40,45,49,56,59,65,72,73,81,82,86,94,94, 97,104,106,114,116,118,119,121,123,125,135,138,140,156–158,163,175,176^ (online supplemental appendix E; Table E2).

Various physical demands are required in different performance settings and among different genres and instruments played, and may influence how JH affects musicians. Most sources from both lists included musicians from professional,^3,28,43,46,59,94,98,102,103, 108,109,116,128,132,135,136,140,156,157,163,165,166,175,176,186,186^ higher education,^3,43,46,52,59,61,63,71,77,80, 84,100–102,107–110,113,122,142,144,151,156,166,185^ and classical ^3,8,28,50,80,98,102,107,109,110,135,136,138,140, 151,156,157,163,165,166,175,176,186^ settings, and fewer than 10% of sources included settings with primary or secondary school students^8,41,50,68,93,99^ or non-classical^94,104,151,172,186^ and amateur musicians^27,43,46,59,108,109,128,184^ (online supplemental appendix E; tables E3-4).

### Publication types and narrative or research types

To better understand the types of sources that comprise the collection of literature on JH, information on publication and study type was charted. First, sources in L1, L2, and L3 were categorized by publication type. In a separate categorization, sources from each list were grouped by narrative or research study and then consolidated into broader categories for more efficient comparison between lists.

Publication type was charted using the following categories:

- **Academic journal articles:** from peer-reviewed journals (L1, *n =* 129; L2, *n =* 56; L3, *n =* 18)
- **Magazine articles:** from consumer and trade magazines (L1, *n =* 16; L2, *n =* 13; L3; *n =* 9)
- **Books:** books, textbooks, and manuals (L1, *n =* 16; L2, *n =* 7; L3, *n =* 1)
- **Academic papers:** theses and dissertations (L1, *n =* 4; L2, *n =* 2; L3, *n =* 2)

Most sources from all lists were published in academic journals (78%). However, L2 has a higher ratio of academic articles compared with magazine articles than L3.

Data on narrative and research study types were charted to identify gaps in literature (figure 4). L1 was not charted because it contains duplicate data. Data from L3 is presented despite containing some duplicate data to provide insight on details of JH-focused sources. Narratives and research study types were first grouped corresponding to JBI critical appraisal tools^21^, then condensed into the following categories:

- **Narrative:** expert opinions (music and medical professionals) and historical narratives
- **Qualitative Study:** case reports and large-scale qualitative studies
- **Quantitative Study:** randomized controlled trials, and case-control, cross-sectional, cohort, and prevalence studies The most common source type in both lists was expert opinion.^26,27,29,35–38,40,45,45,49,52, 56,59,61,65,68,72,73,77,81,82,86,94,97,99,100,104,106,109,113,114,118,119,121,125,132,138,140,158,175,176,186^ Most quantitative studies ^3,8,50,71,107,108,110,116,122,123,128,135,136,142,144,157,171,185^ were prevalence studies.^28,38,39,41,43,46,57,62,63,79,93,101,132,151,156,163,165,166,172,184^ One randomized controlled trial^151^ was conducted among L2 sources, but no other studies in which causal relationships can be established were conducted. The ratio of research studies to narratives in L2 (about 4:1), is much higher than that of L3 (about 2:1).

**Figure 4.**
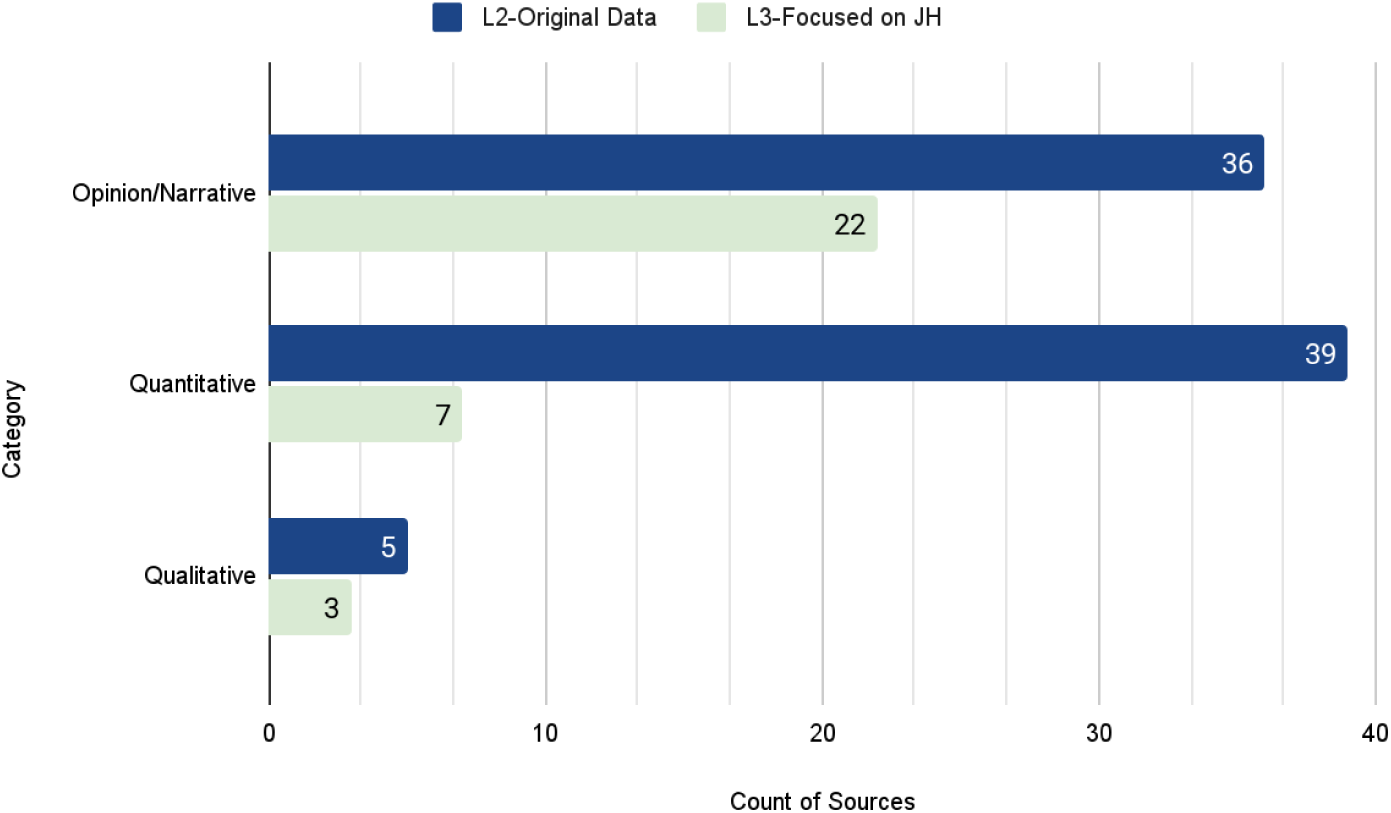
Type of Study or Narrative

### Relationship between hypermobility and injury

To answer Question 1, “What is known from the literature about the relationship between hypermobility and playing-related injury and difficulty among instrumental musicians?” the extent to which existing information on the relationship between JH and PRMPs is supported by research or expert opinion rather than general or unsubstantiated claims, relevant data were collected. Sources containing original conclusions about this relationship were identified and categorized based on whether conclusions were from the author’s professional experience or from study findings. A summary is provided in Figure 5, and described below.

**Figure 5.**
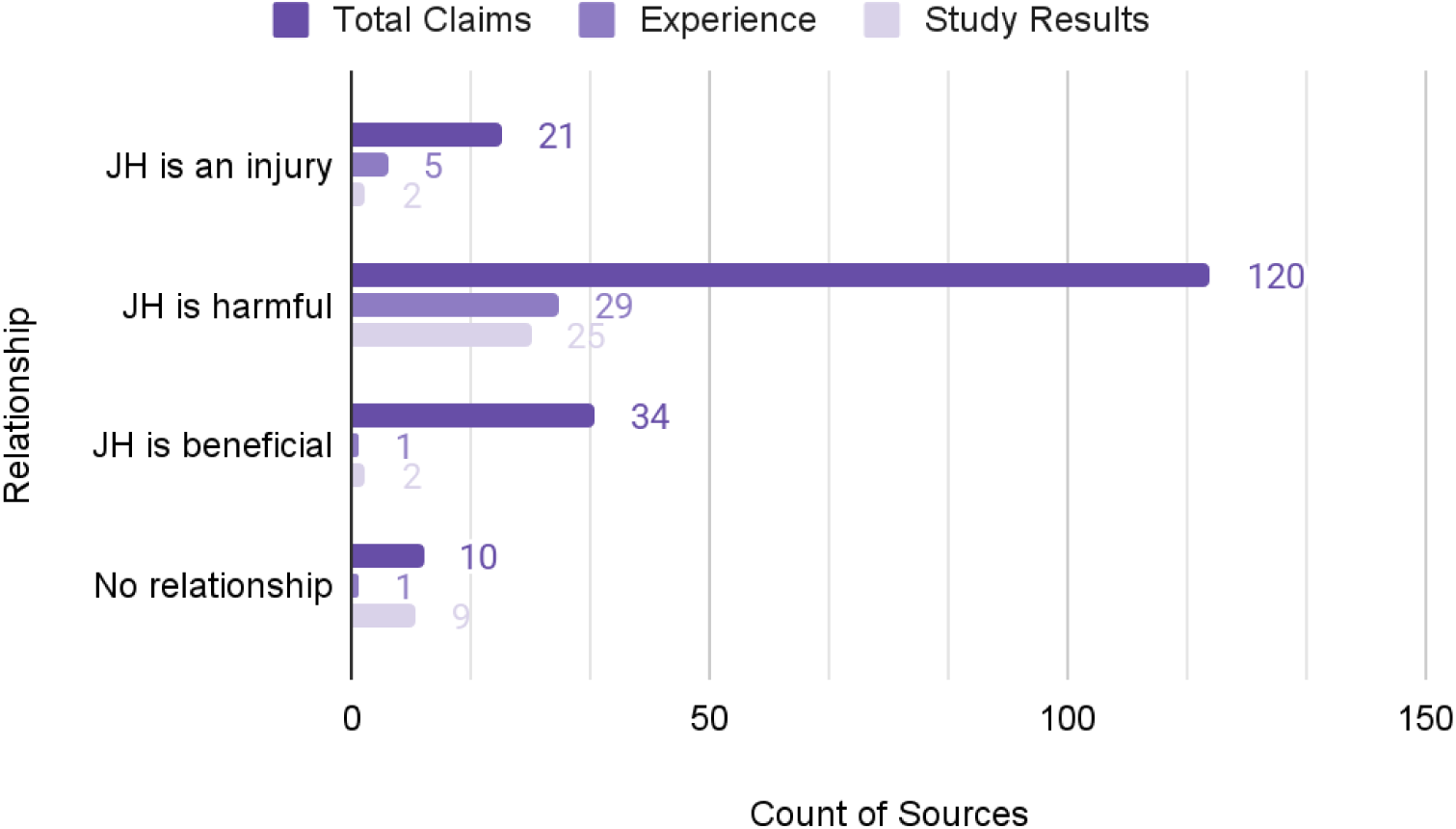
Relationship between JH and PRMPs: Article claims

### Benefits of joint hypermobility in musicians

About 20% of review sources contained assertions that JH may be an advantage for musicians. ^3,27,30,32,36,41,47,54,55,58,62,63,72,90,92,95,97,109,114,115,118,120,124,125,127,129,131,137,139,140,174–176,187^ This claim was only supported by original findings from two studies^3,62^ and the expert opinion of one author^114^ in L2. Some assertions came from authors who referenced famous musicians such as Niccolò Paganini, Franz Liszt, and Sergei Rachmaninoff, whose JH may have contributed to their ability to reach difficult positions on their instruments,^62,109^ as is evident in the extreme finger spacing and hand positions required to perform their compositions.^114^ Larsson *et al*.^62^ found that two of 19 flute players with musculoskeletal symptoms had hypermobility, and nine of 13 flute players who did not have symptoms had JH. They concluded hyperlaxity may be protective against injury for situations requiring non-weight-bearing fast movements. Burkholder and Brandfonbrener^41^ suggested ligament laxity may be protective against carpal tunnel syndrome because females had higher rates of ligament laxity and lower rates of carpal tunnel syndrome than males. Winspur and Wynn Parry^131^ and Dawson^45^ noted JH in fingers may allow guitarists to reach difficult chords easily. Similarly, Beighton^109^ hypothesized ligament laxity may enable violin players to achieve large reaches across the fingerboard. Violin expert Mimi Zweig suggested hypermobile finger joints can help violinists achieve a rich vibrato.^27^

### Disadvantages of joint hypermobility in musicians

In contrast, authors from over 70% of sources concluded JH is harmful and was supported by both findings from original studies ^8,35,39,40,46,50,62,80,84,93,98,108,110,116,123,125,128, 142,151,157,171,184^ and personal experiences of source authors. ^27,29,36,37,45,49,52,59,65,68,72,77,81, 82,86,94,97,99,100,106,109,113,114,116,118,119,121,125,186^ This assertion was based primarily on two observations. First, researchers within the medical field reported that females exhibited higher rates of both JH and injury compared to males.^61^ Second, medical practitioners noted that a significant number of injured musician patients were hypermobile, leading them to suspect a connection between JH and injury.^35,110^

Two hypotheses emerged from these observations. One was that playing an instrument causes JH, an observation made in 12% of review sources and supported by findings from two studies^108,165^ and personal experiences recorded in five sources.^36,37,63,73,114^ The second was that inherent JH increases the risk of injury while playing a music instrument. To investigate these observations further, lists of review sources containing original data comparing JH and injury prevalence between males and females, hypermobile and non-hypermobile musicians, and between musicians and the general population were compiled (online supplemental appendix D, table D4).

Review sources containing original biological sex-based prevalence data (*n =* 15) were compiled. In agreement with medical research, all but one author from this list reported higher JH prevalence in females than males,^28,61,62,107,110^ nine of which also confirmed higher injury prevalence among females.^8,35,39,41,63,83,128,136,172^ One source showed no difference between males and females;^165^ however, over 16% of participants were missing at least one component of the JH assessment, which may have influenced results.

Review sources containing original prevalence data on injury in injured compared to non-injured musicians (*n =* 21) were compiled. Of these, researchers from eight sources reported that injured musicians had higher JH prevalence than their uninjured counterparts.^8,35,36,39,41,62,93,101^ Authors of two sources found no difference between groups;^71,184^ however, Lonsdale *et al*.^184^ noted injured hypermobile participants reported that their problems were directly caused by JH. The remaining 11 sources contained relevant correlational information, but authors did not make a determination regarding injury risk.^38,63,79,107,110,125,132,136,156,163,165^

To test the hypothesis that playing a musical instrument causes JH, researchers compared JH prevalence among musicians to that of the general population. To compare related data and conclusions, research studies containing original prevalence data on JH in musicians (*n =* 20) were compiled. Among identified sources, 12 contained data but no conclusions.^8,36,39,61,71,79,101,125,156,163,165,184^ Among those that did, conflicting conclusions were found. Researchers from two sources found no difference in JH rates between musicians and non-musicians,^122,123^ however, Miller *et al*.^123^ reported that hypermobile musicians experienced more JH-related pain than non-musicians. In contrast, two sources contained instances wherein guitar players^110^ and piano players^132^ had less wrist mobility than the general population. Authors of four sources reported higher rates of JH in musicians than the general population.^35,41,93,108^ Although Brandfonbrener suspected there was genetic influence for some hypermobile individuals, she also noted that woodwind players accounted for 20% of her clients and nearly always exhibited thumb hyperlaxity.^39^ She suggested ligament damage due to bearing the weight of the instrument caused the laxity.

Authors of personal experiences and historical narratives were supportive of Brandfonbrener conclusion.^30,73^ Additionally, O’Shea proposed that Paganini’s JH may have been exacerbated by his intensive childhood practice regimen through stretching connective tissue before ossification occurred.^124^ More recently, *School Band & Orchestra* published an article in which a chiropractic doctor recommended wrapping joints while playing to prevent JH, implying that playing an instrument may contribute to joint instability.^68^

### Negative impacts of joint hypermobility on musician health and performance

Specific **health impacts** mentioned in sources were charted and are listed below (table 1). Negative impacts of JH on instrument **performance impacts** mentioned include joint instability, ^8,26,27,30–32,35–40,42,45,47,49,50,52,54,55,59,62,65,68,72,73,80,81,83,84,87–90,92,94–97,99,100,104,106,109, 112,114–116,116,118–120,123,127–130,168,187^ altered movement patterns, ^8,27,31,37,38,40,42,45,47,52,55,59,65, 68,72–74,77,80,81,84,89,90,96,97,100,106,112,114,116,118,121,123,126–128,187,188^ compensation by surrounding muscles, ^8,27,30,32,37,38,40,54,55,62,72,80,81,83,87,88,90,96,106,109,112,114,120,123,127,128,130,168^, impaired dexterity, ^8,27,30,45,52,62,68,77,80,82,84,89,90,96,97,99,100,106,115,120,121,184^ fatigue,^8,27,30–32,42, 54,62,72,74,77,84,87,94,120,130,130,186,188,189^ excessive tension, ^8,27,30,37,38,40,45,68,77,84,90,96,100,106,115, 123^ difficulty supporting the instrument,^27,31,35,36,45,55,65,68,80,89,92,96,97,115,120,123^ impaired proprioception,^8,27,38,42,45,72,81,84,108,114,116,126,127,129,131^ altered posture, ^31,38,40,45,52,65,72,77,89, 114,116^ excessive pressure, ^8,27,38,68,77,100,115,129,189^ limited technical achievement,^31,47,52,68, 73,80,120,157,186^ reduced strength,^115,121,130,131,188^ spontaneous dislocation,^27,89,95^ impaired tone,^99,100^ impaired focus,^72^ inhibited breathing,^77^ and difficulty sitting^31^.

**Table 1.** Health Impacts of Joint Hypermobility in Musicians.

| Category | Condition |
| --- | --- |
| **Pain** | **Pain** |
| **Bone & Joint Issues**<br>27,35,38,55,77,80,84,89,95–98,106,112,114,115,119,120,127,157,188 | Arthritis (9), Osteoarthritis (6), Dislocation (10), Subluxation (5), Misalignment (1), Hip dysplasia (1), Increased hypermobility (4), Bleeding in joint spaces (1), Swan neck (1), TMJ disorders (8), Spinal complications (6), |
| **Muscle & Soft Tissue**<br>8,27,30–32,35,38,40–42,45,46,54,55,62,68,72,77,81,87–90,96,97,109,112,114–116,119,120,123,127–131,168,188,189 | Stressed muscles (37), Muscle imbalance (11), Spasm (5), Weakness (3), Weak hand intrinsics (1), Flat foot arches (1), Rapid deconditioning (2) |
| **Tendons & Ligaments**<br>10,30,32,35,36,38,39,41,53–55,77,80,84,87,89,92,109,110,112,115,118–120,125,127,129–131,182,189 | Rheumatological complaints (14), Sprains/Strains (7), Ligament rupture (1), Tendinitis (8), Collagen stiffening (1), Tendon swelling (17), Ganglion cysts (1), Joint effusion (1), Fibromyalgia/Myofascial pain (6), Trigger points (1) |
| **Neurological Issues**<br>32,36,39,54,55,80,84,87,89,95,97,114,120,127,182,188 | Dystonia (2), Impaired reflexes (1), Traction damage (4), Nerve compression (9), Nerve entrapment (5), Neuropathy (3) |
| **Mental Health**<br>72,84,114 | Acute Stress Response (2), Other difficulties (3) |
| **Functional Issues**<br>30,31,35,37,38,50,63,67,72,84,89,92,99,109,112,115,118,119,123,126,188,189 | Decreased range of motion (4), Deformity (4), Disability (8), Acute injury (2), Minor trauma (13), Headaches (1), Autonomic dysfunction (1) |
| **Systemic / Visceral**<br>31,72,84,97,131 | Breathing difficulty (1), Digestive issues, (2), Heart complications (1), Uterine/rectal prolapse (1), Varicose veins (1), Vascular compression (1), Visual disturbances (1) |
| **Clinical Consideration**<br>89,97,127 | Not all have symptoms |

### At-risk populations

To answer Question 2, “Are there subpopulations of instrumental musicians with an elevated risk of playing-related injury and difficulty due to hypermobility?” information related to populations with connections to or elevated levels of JH was collected from review sources. Females were identified most frequently as having high rates of JH or risk of injury,^3,28–36,38,39,41,45,46,50,53,61–63,67,78,79,84,89,95,105,107–109,114,115,118–120,126–128,131,134,136, 148,155,156,159–161,166,168,173,181,183,190^ followed by children under age 18.^8,35,41,45,55,62,68,83,93,96, 114,119,127,166–168^ Populations of certain races and ethnicities mentioned included Asians, ^39,39,62,99,118,127,131^ Africans,^62,89,99,114,118,127^ Indians,^89,114,118,127^ Iraqis,^114^ and Middle Easterners.^99^ Instrumentalists mentioned were string players^77,149,180,189^ and keyboard players.^77,180^ Other populations mentioned were those who are tall and thin,^119,127,131^ and those who were trained in dance in childhood.^114^ One author mentioned individuals with developmental disorders such as ADHD and autism are more likely to be hypermobile than the general population.^72^

### Injury prevention and supportive strategies

To address Question 3, “What supportive methods have been used for instrumental musicians with hypermobility?” techniques reported in the literature were noted and compiled (supplemental online appendix D; table D5). These included physiotherapy,^8,27, 30,38,42,45,47–49,52,54,55,59,72,80,81,84,89,90,92,96,97,114–116,120,125–127,129,131,168,186,191^ orthotics, ^8,27,30,35, 38,39,42,47,48,50,54,55,59,72,80,84,89,90,92,94,96,109,115,116,120,127,168, 187–189^ instrument technique adaptations, ^8,35,37–39,45,55,62,72,77,80,81,84,89,96,97,100,106,115,116,119, 121,127,186,189,191^ modifications to practice habits and/or repertoire, ^35,37–39, 45, 50,55,62,66,72,77,84, 89,94,106,109,115, 116,119,166,186^ patient education, ^8,27,35,37–39,50,55,72,80,84,89,90,102,106,115,116,125,129, 189,191^ instrument adaptation, ^27,37,38,55,66,72,80,89,92,95,96,116,191^ early diagnosis,^26,35,37,38,50,52,80, 81,89,96,97,120,131^ surgery,^30,42,50,54,89,92,96,112,116,188^ supportive taping,^27,39,50,55,84,90,96,127^ rest,^27,50,72,73,89,109, 131,186^ medications,^89,109,110,126^ hypermobility screening,^35,55,191^ modifying life activities,^38,90,116^ switching instruments,^62^ massage,^55^ and authors of three sources mentioned surgery was not recommended.^45,115,116^

### Reported weaknesses in research

Development of future research studies requires understanding weaknesses and gaps identified by those with experience with the topic. Information was collected on weaknesses in research identified by L1 source authors. Authors mentioned problems with JH measurement standards most frequently,^31,35,37,38,44,55,61,62,84,89,93,108,114,123,136,146, 160,162^ and many mentioned that further study is needed.^31,35–37,39,50,58,62,63,92,93,108,118,123, 128,141,143^ Additional research gaps included missing populations and lack of longitudinal studies,^8,32,35,39,41,89,110,162^ conflicting data or statements and inaccurate analyses,^37,38,58, 100,104,114,162,183,191^ issues with research methods such as small sample size, recall bias, missing information, low response rates, inability to generalize to a larger population, and inadequate statistical analysis,^8,31,35,41,50,62,89,136,143,162,163^ and not enough information or focus on hypermobility.^35,62,77,80,84,89^ To identify whether reported weaknesses from earlier studies have been addressed, sources published after 2010 were examined; they contained similar reports of weaknesses.

### Methods of measurement

To address identified gaps, methodological problems, and limitations of measurement tools in JH research, practices within existing research need to be understood. To this end, data were collected on JH measurement procedures used in quantitative and qualitative studies from L2. The complete data set for all lists can be accessed at https://osf.io/6jynk/(15). JH measurement methods were grouped into the following categories:

- **Beighton:** derivatives of the Carter-Wilkinson scale such as Beighton and Brighton Scales
- **Individual:** assessing specific joints with instruments such as a goniometer or Leeds hyperextensometer
- **Hybrid:** a combination of Beighton and Individual joint assessment
- **Self-report:** participant-identified hypermobility, often through surveys or asking if they call themselves “double jointed”

In 27% of research studies (*n* = 41) a derivative of the Beighton Scoring System was used to measure hypermobility,^3,8,46,50,62,63,107,128,136,142,165,185^, in 14% individual joints were measured,^28,38,39,41,122,157^ in 6% a hybrid of Beighton and individual joint measurement was used,^84,108,110,123,132,151^ in 7% self-report was used,^71,93,184^ and in 36% of studies measurement tools were either not specified or not measured.^43,57,79,80,98,101, 102,116,135,144,156,163,166,171,172^

### Influences on joint laxity

Some review authors noted factors that may influence joint laxity, which could impact both measurement procedures and the interpretation of JH assessments. Information on these influencing factors was extracted and summarized. Reported participant characteristics influencing laxity included:

- **Biological factors:** sex,^29–31,33–36,38,39,41,50,53,54,61–63,67,79,84,89,90,95,107,109,110,114,118, 126–128,131,148,155,183^ age,^35,38,39,41,50,55,61,62,89,96,107,109,114,119,127–129,131,160^, race and ethnicity, ^30,38,39,62,89,107,114,118,127,131,160^ dominant hand,^89,114^ bone contour,^77,114^ collagen structure,^77,114^ muscle tone,^77,114^ and proprioception^77^
- **Hormonal influences:** menstrual cycle,^89,114,126^ pregnancy, menopause, breastfeeding, contraceptive use, and gynecological conditions^114^
- **Environmental and contextual factors:** joint trauma,^26,30,35,37,88,89,96,188,189^ repetitive or extreme loading,^35,37,89,95,99,104,108,188,189^ time of day,^127,131^ and weather conditions.^89^

These variables were mentioned in the context of their potential to cause localized or generalized increase in laxity. However, few researchers reported considering these factors when designing studies.

## Discussion

A definitive answer was not found to answer the research question, “What is known from the literature about the relationship between hypermobility and playing-related injury and difficulty among instrumental musicians?”. Discussion on joint hypermobility among musicians was scarce, and only half of the sources consisted of research studies. Correlational and anecdotal data were indicative of a positive relationship, however, research studies contained methodological limitations.

The answer to Question 2, “Are there subpopulations of instrumental musicians with an elevated risk of playing-related injury and difficulty due to hypermobility?” appears to be affirmative and include individuals affected by female hormone cycles, children, individuals not of European descent, and neurodivergent individuals, although a lack of empirical studies among suspected groups prevents confirmation of this assertion.

Question 3, “What supportive methods have been used for instrumental musicians with hypermobility?” has been addressed. Various versions of physiotherapy, orthotics, adapting instruments, and changing practice routines or life habits were recommended. Some authors suggested surgeries while others warned against them. Early screening and diagnosis and patient education on joint protection were recommended. However, many authors reported on their experience rather than empirical studies, and many were not credentialed medical practitioners. Further, limitations in the quality and scope of research studies restrict what conclusions can be drawn.

### Research gaps

The small percentage of JH-focused sources and high ratio of anecdotal reports compared with sources focused on broader PRMPs are indicative that more research is needed on musicians with JH. Further, most research on musician injuries, including research on JH, has been focused on prevalence. To determine which treatments and teaching practices are safe and effective, longitudinal studies and randomized controlled trials are needed.^169,192^ Although researchers in the field of performing arts medicine are increasingly pursuing these study types,^170^ more studies should include JH as a measured variable, even when it is not the primary focus, to help clarify JH’s role as a potential risk factor.

Significant gaps in research were found among vulnerable populations. Females were more likely to be hypermobile than males and were more likely to have injuries. Female hormone cycles are known to have an impact on laxity but were not considered by many researchers.^50,109,114^ This could have further implications for populations influenced by hormone cycles, such as transgender individuals,^193,194^ who were not identified in review sources. Children were not included in most studies but have higher JH prevalence and similar injury prevalence as adult musicians.^8,109,127,130^ Individuals of European descent had the lowest JH prevalence but most studies were conducted among these populations, and few studies included individuals not of European descent.^114^ Amateur musicians were not included in many studies but may be exposed to significant injury risk-factors.^195,196^ There is a connection between JH and neurodivergent individuals such as those with autism and ADHD,^197–199^ but only one source included this as a point of discussion.^72^

Neurodivergent populations may be particularly vulnerable due to barriers to accessing care. Communication challenges and difficulties with planning and scheduling can make it harder to seek medical support or follow through with injury prevention and treatment.^200,201^ These challenges may also contribute to underrepresentation in research. Neurodivergent individuals are more likely to experience both acute and chronic musculoskeletal injuries than the general population,^202^ and JH itself is classified as a musculoskeletal disorder.^203^ However, some review sources excluded individuals who had been previously diagnosed with a musculoskeletal disorder,^60,117,157,172,185^ potentially limiting inclusion of this population. Several researchers also reported data loss due to missed appointments^66,151^ or incomplete paperwork,^165,172^ which may disproportionately affect neurodivergent participants.

### Inconsistent conclusions and results

Inconsistencies among study results, and conclusions are indicative that there may be a disconnect between research, and the lived experience of musicians with JH. A claim that hypermobility may be beneficial for musicians was highly prevalent among review sources but only supported by findings from two studies.^3,62^ Clinicians such as Brandfonbrener criticized these findings citing the high rates of hypermobility among injured musicians who visited her clinic. She further noted that observing musicians with JH while playing their instruments was important in understanding the effects of the interaction.^37^ The claim that JH is harmful for musicians was well supported by both scientific studies and expert opinion, but several source authors reported conflicting data showing there was no relationship between JH and injury.^28,71,107,122,132,136,166,185^ Conflicting results were also found among studies comparing injury rates between musicians with and without JH. Further, incongruence between results and participants’ experience was noted by Lonsdale *et al*. who mentioned that although no differences were found in injury rates between groups, injured participants with JH felt their problems were directly caused by JH.^184^ Only a limited number of sources contained prevalence data comparing JH musicians to the general population, there were conflicting results among them, with researchers finding a loss of mobility in some joints of musicians,^110,132^ while others found no difference^122,123^ or higher rates.^35,41,93,108^ Although Miller *et al*.,^123^ found no difference in JH rates between musicians and non-musicians, they noted hypermobile musicians experienced more JH-related pain than non-musicians. These inconsistencies and incongruences illustrate possible methodological weaknesses and a need for better understanding of the dynamics affecting musicians with JH.

### Knowledge limitations among stakeholders

Knowledge limitations related to JH and the requirements of instrument performance may contribute to inconsistencies and incongruencies in study results and author conclusions. Many medical practitioners, researchers and musicians do not receive specific training on JH and may not fully understand its implications.^204,205^ This phenomenon is illustrated by examples of injured musicians from review sources who were misdiagnosed for years prior to being diagnosed with JH^102,110^ and music teachers asking questions about joint collapse among their students.^27,106^ Some source authors noted factors that influence joint laxity such as dominant hand, hormonal influences, and weather conditions, but few researchers considered these factors when assessing JH.

In addition to a lack of specific training on JH, few medical practitioners receive specific training on the demands of instrument performance^89,163,206^ and may misinterpret performance-related data as a result. For instance, Larsson *et al*.^62^ concluded JH was helpful for joints requiring repetitive motion such as wrists and fingers of flute and violin players, and harmful for weight-bearing joints because they found fewer hand and wrist injuries in flute players with hypermobile wrists than those without. However, the conclusion may be in error because fingers and wrists of flute and violin players are assistive in supporting the instrument and must bear pressure at times, and several authors specifically mentioned flute and violin players experienced PRMPs related to their JH.^80,102,191^ These knowledge gaps may have influenced research design, interpretation, and JH measurement and contributed to the conflicting results and conclusions found in the scoping review.

### Limitations of research diagnostic criteria and measurement tools for joint hypermobility

Inconsistencies and incongruencies in study results and conclusions among review authors may also be influenced by weaknesses in diagnostic criteria and measurement tools for JH, as noted by several review sources. Most researchers used a version of the nine-point Beighton Scoring System, to assess JH,^207^ but the cutoff scores used to diagnose JH vary widely, typically ranging from 4 to 7 hypermobile joints.^50,208^ However, some researchers suggested that even a score of 1 can be clinically relevant for musicians.^62,89,127,188^ Although the Carter-Wilkenson, Beighton, and Brighton Scoring Systems are similar, each includes slight variations, and researchers sometimes modified them further by adding or omitting measurements of certain joints. These inconsistencies may help explain why reported JH prevalence in the general population ranges from 5% to 34%.^7^ Without standardized diagnostic criteria and measurement tools, it is difficult to determine whether musicians truly experience higher rates of JH than the general population.

In addition to the variability in cutoff scores, other limitations exist in scoring systems used in review sources. Beighton-based systems present specific weaknesses. Bird, who co-authored “Hypermobility of Joints” with Beighton explained that this scale was developed to conduct evaluations as quickly and reliably as possible for large scale studies conducted in South Africa and were not intended for pathological evaluation.^114,207^ Although the Beighton Scoring System has a 99.3% specificity rating, meaning that a positive test is highly accurate, its sensitivity rating is only 13.8%.^207^ This low sensitivity means many hypermobile individuals may receive false-negative results and be overlooked.

The Beighton Scoring System, adapted from the similar Carter-Wilkinson Scoring System, includes one-directional measurement of nine joints including wrist flexion, pinky extension, elbow extension, lower back flexion, and knee extension.^109^ However, there are approximately 350 joints in the human body and many joints that are particularly relevant to musicians, such as those in the fingers and shoulders, are omitted in the Beighton Scoring System.^37^ Researchers recognizing these limitations expanded their assessments to include additional joints.^(35,62^ Shoulder instability is also a common issue among both musicians^209^ and hypermobile individuals,^193^ yet is not considered in the Beighton scoring system.^207^ Given the unique physical demands of playing musical instruments, JH assessment tools for musicians should be revised to accurately reflect the joints and movements most impacted by instrument performance. The Upper Limb Hypermobility Assessment Tool (ULHAT) developed in 2018, intended for use in occupational populations, has higher sensitivity and specificity scores than the Beighton scoring system. Although it has not been validated for all populations, it has potential for use in screening musicians for JH.^208^

### Implications for research

Research on hypermobility among instrumental musicians can be improved and expanded. Researchers conducting quantitative studies should prioritize longitudinal studies and randomized controlled trials to establish causal relationships and evaluate effective supportive strategies. Studies focusing specifically on hormonal influences and at-risk populations should be conducted. Researchers should include JH measurements, descriptions of methods and definitions, and make them publicly available, even when the project is not specifically focused on JH. Although logistical challenges may prevent measuring all subjects under the same conditions such as time of day, warm-up status, or weather patterns, these factors could be noted to aid data analysis. Additionally, researchers should consult and collaborate with experienced musicians when designing and analyzing research related to musicians to develop an accurate understanding of the physical demands of playing instruments.

Further, qualitative research is needed to advance understanding of JH among musicians. Recruiting sufficiently large samples for longitudinal or randomized controlled trials, especially when analyzing subgroups by instrument type, remains challenging. Moreover, assessing individual joints is often impractical, leading to the use of less-specific tools and limiting the applicability of multivariate analyses, as noted by Larsson *et al*.^62^ Researchers in performing arts medicine are increasingly advocating for individualized approaches to both research and treatment.^182,210,211^ Many review authors included personal experiences and case studies which provided relevant information not available through controlled trials, and in some cases, contained information that contradicted study results. These perspectives, along with the low sensitivity rate of the Beighton Scale, are indicative that more sensitive tests are needed to assess joint hypermobility among musicians. Consequently, qualitative studies should be conducted involving a) lived experiences of musicians with hypermobility, b) music teachers who work with hypermobile students, and c) medical practitioners who treat hypermobile musicians. Thematic coding of such data could allow researchers to evaluate joint use by specific instrument, performance and health effects, and relevant lifestyle factors. These insights could support the development of more accurate and practical measurement approaches suitable for large-scale quantitative research.

## Conclusions

Musicians with Joint Hypermobility (JH) remain underserved across research, pedagogy, and clinical practice. Quantitative research focused on this population needs to be expanded and improved. This includes conducting randomized controlled trials and longitudinal studies, developing more sensitive JH measurement tools tailored to the needs of musicians, and ensuring that relevant data is made publicly accessible.

Researchers should address gaps by including underrepresented populations with elevated JH rates or injury risk such as individuals with hormone-influenced joint laxity, children, non-European racial and ethnic groups, amateur musicians, and neurodivergent individuals. Qualitative research should be conducted to explore the lived experiences of hypermobile musicians, offering nuanced insights that can inform both measurement procedures and broader research design. Additionally, medical training should include more information about hypermobility and important nuances of treating musicians. Training for music educators and musicians should involve health protection, including information about hypermobility. This can help expand understanding of JH and improve treatment, teaching, and research practices.

## Data Availability

The data collected for this study are available in the Open Science Framework (OSF) at https://osf.io/6jynk/. The dataset is licensed under a Creative Commons Attribution 4.0 International License.

## Acknowledgements

This review was conducted as a partial requirement for a Master of Arts degree for M. L. King. King, who is the guarantor for this work, prepared and registered the protocol, conducted the search, conducted screening and discrepancy resolution at all phases of screening, conducted extraction and analysis, and prepared the manuscripts, appendices, and OSF files. Heather M. Macdonald reviewed and approved the Protocol, screened all articles at title/abstract screening, screened many articles at full text, provided input into analysis and manuscript preparation, and provided editing on the final manuscript. Extensive editing and advising was provided by Dr. Jennifer S. McDonel. Additional advising and thesis committee participation was provided by Dr. Sekyung Jang and Dr. David Zuschin. Heather M. Macdonald assisted with the full process of title and abstract screening. Heather M. Macdonald, Mandi Marquardt, Anna-Lisa Davidson, and Jack Browning, assisted with portions of full text screening. Language editing and phrasing clarification were supported by generative AI tools, including ChatGPT^212^ and Gemini.^213^ Table 1 was adapted using assistance from ChatGPT.^212^ References were tracked and added to the text using Zotero 6.0.36.^214^

## HYPERMOBILITY 2025-ONLINE SUPPLEMENTAL APPENDIX

### Appendix A

**Figure A1.**
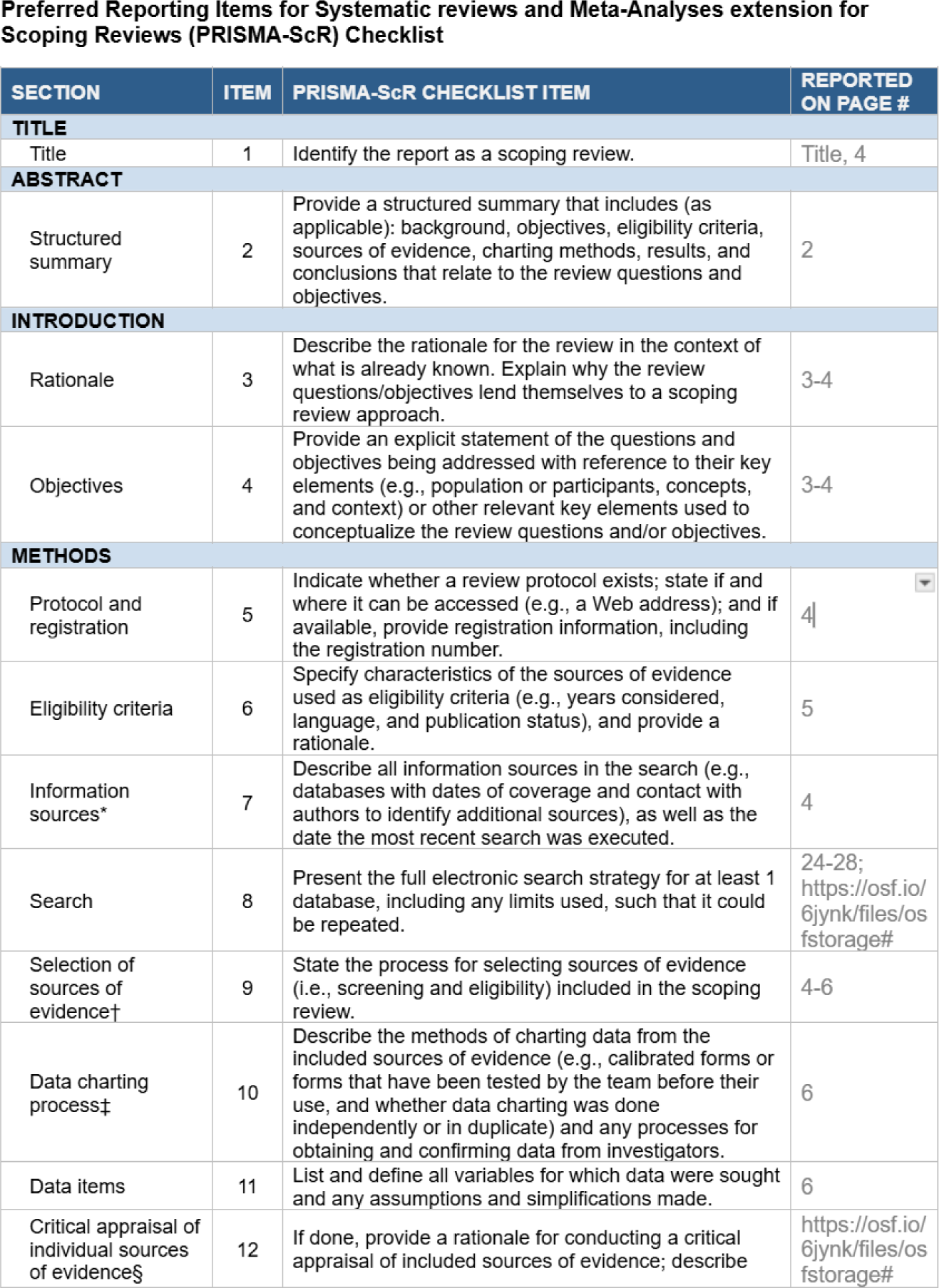
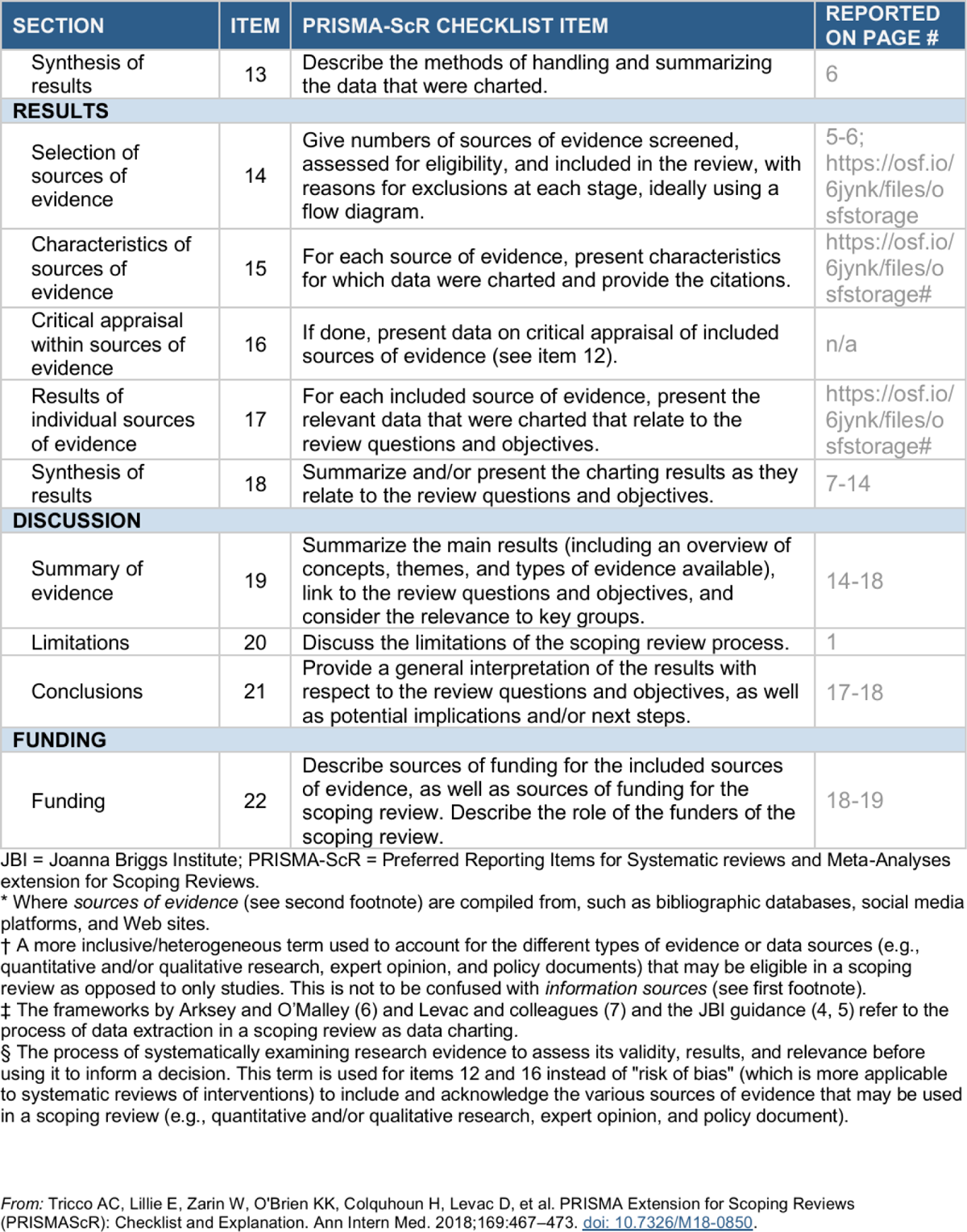
PRISMA-Scr Checklist

### Appendix B

#### Official Search Strategy: Search Query Strings

**Table B1.**
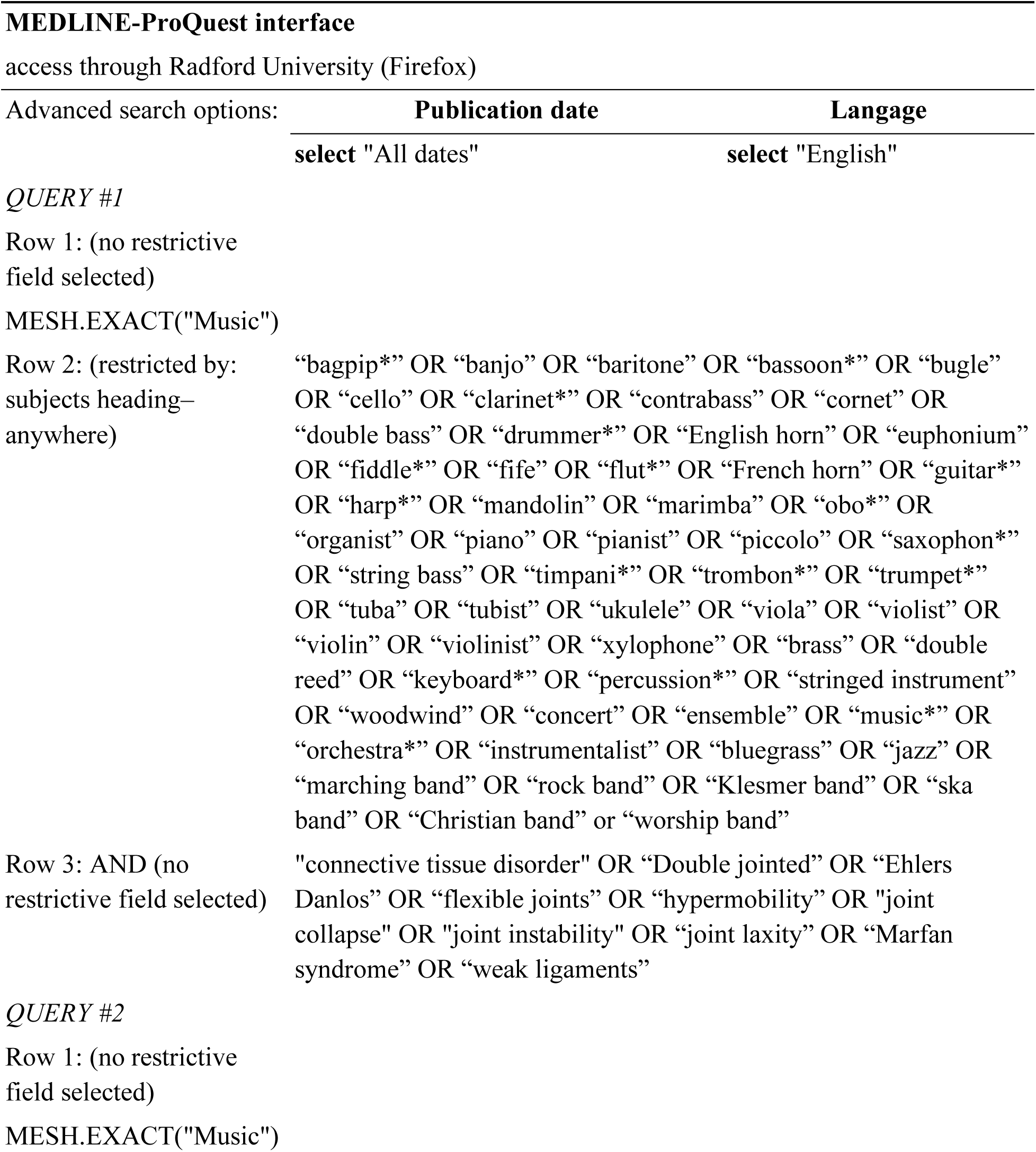
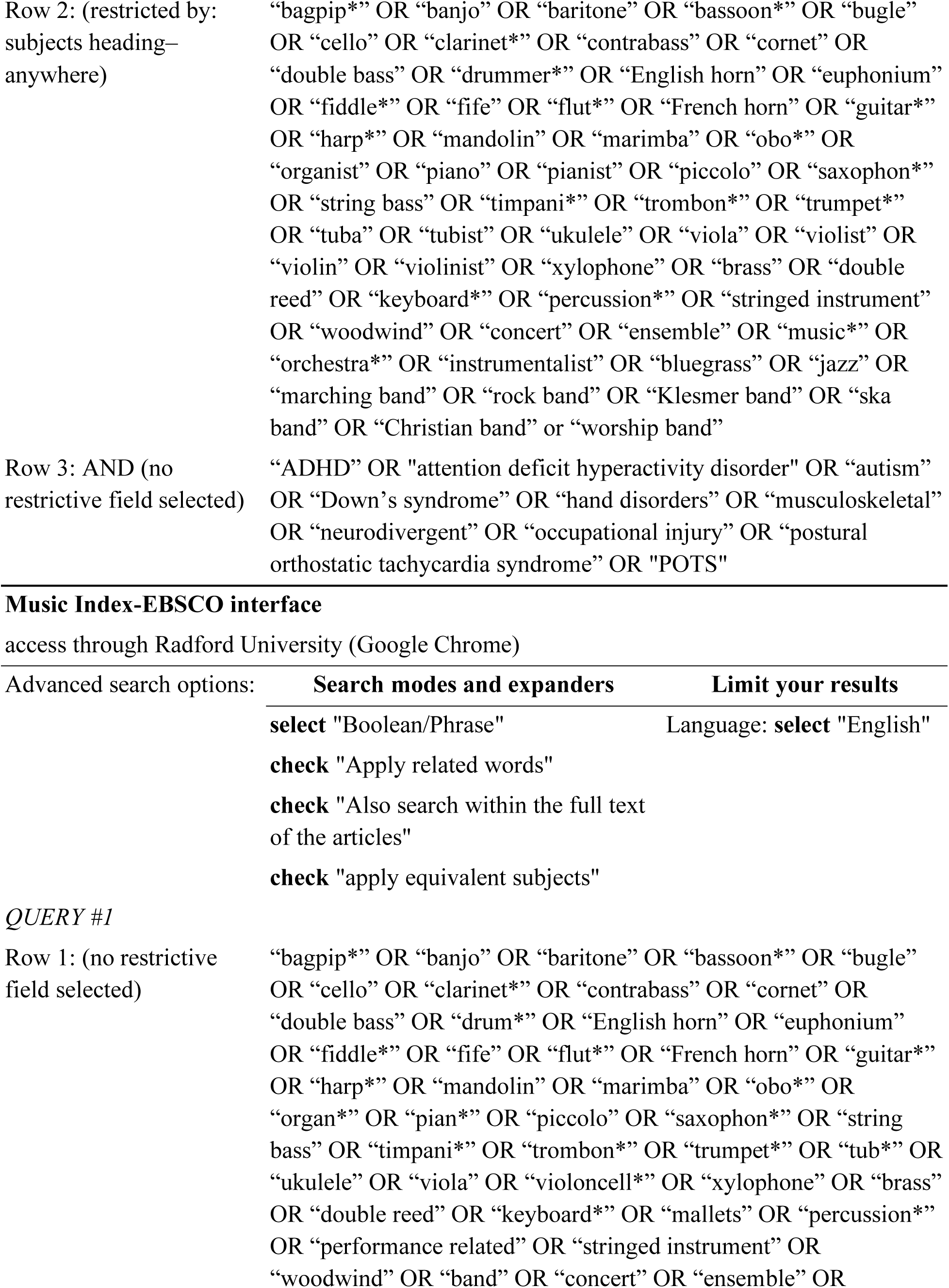
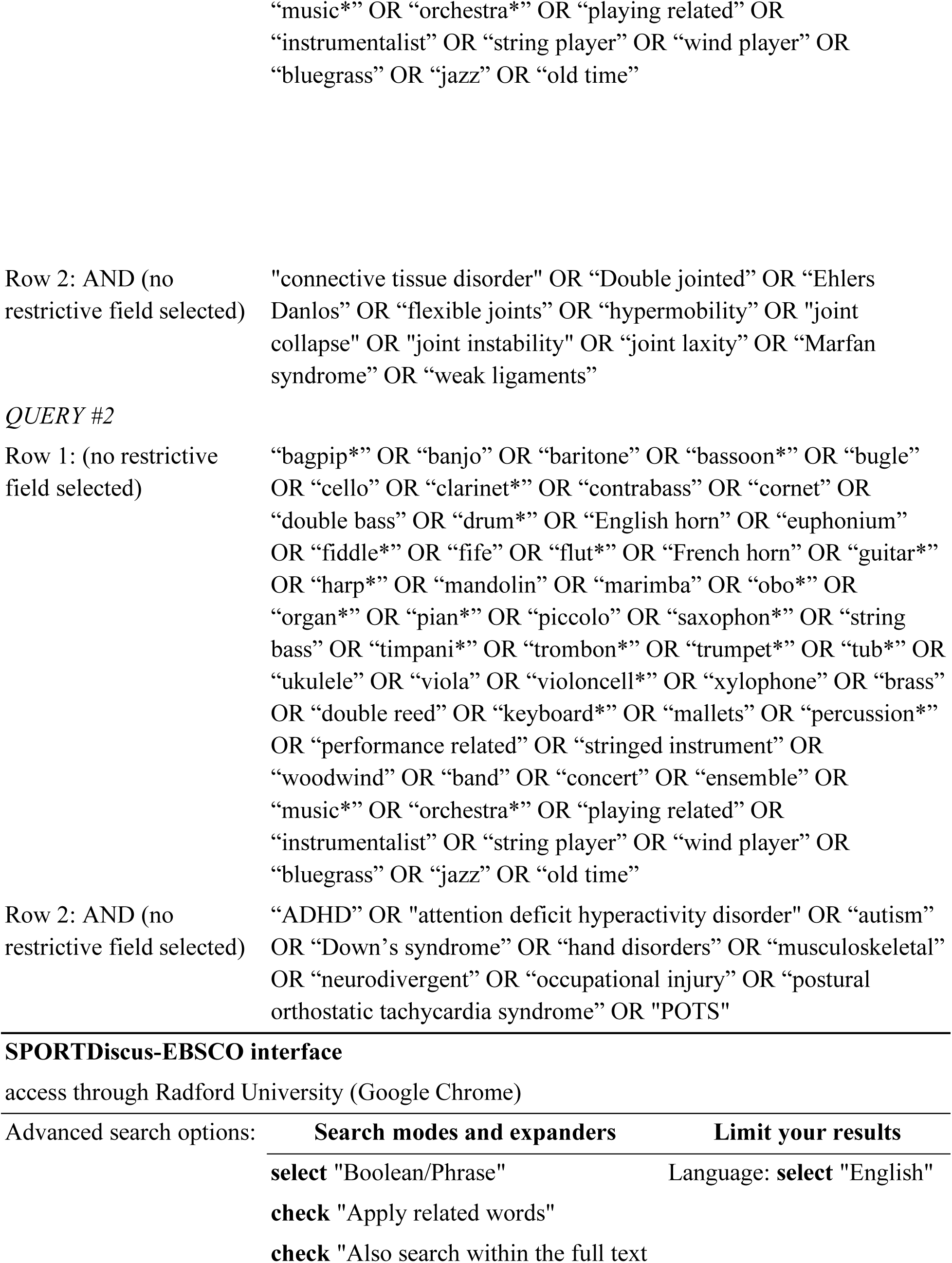
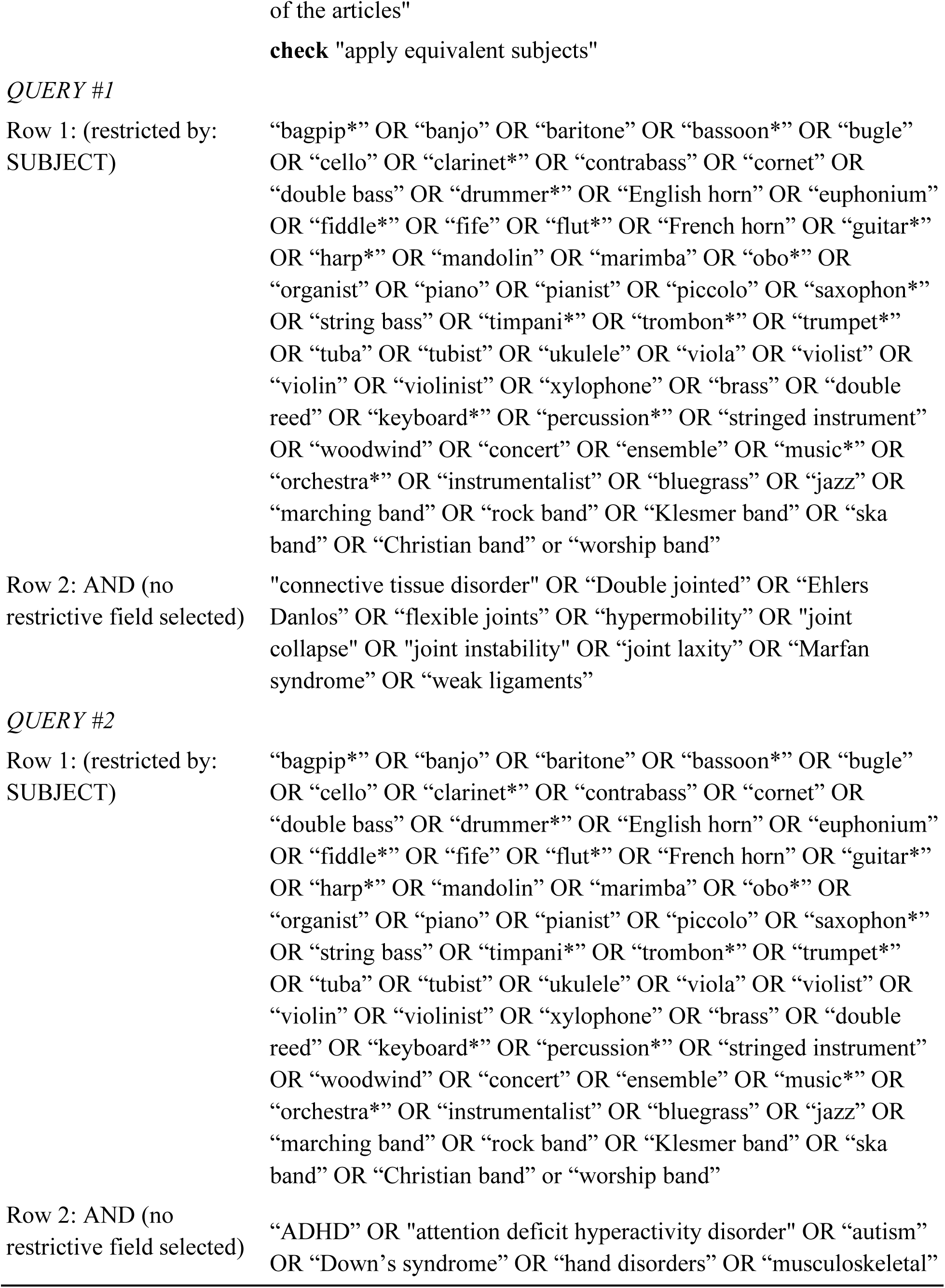
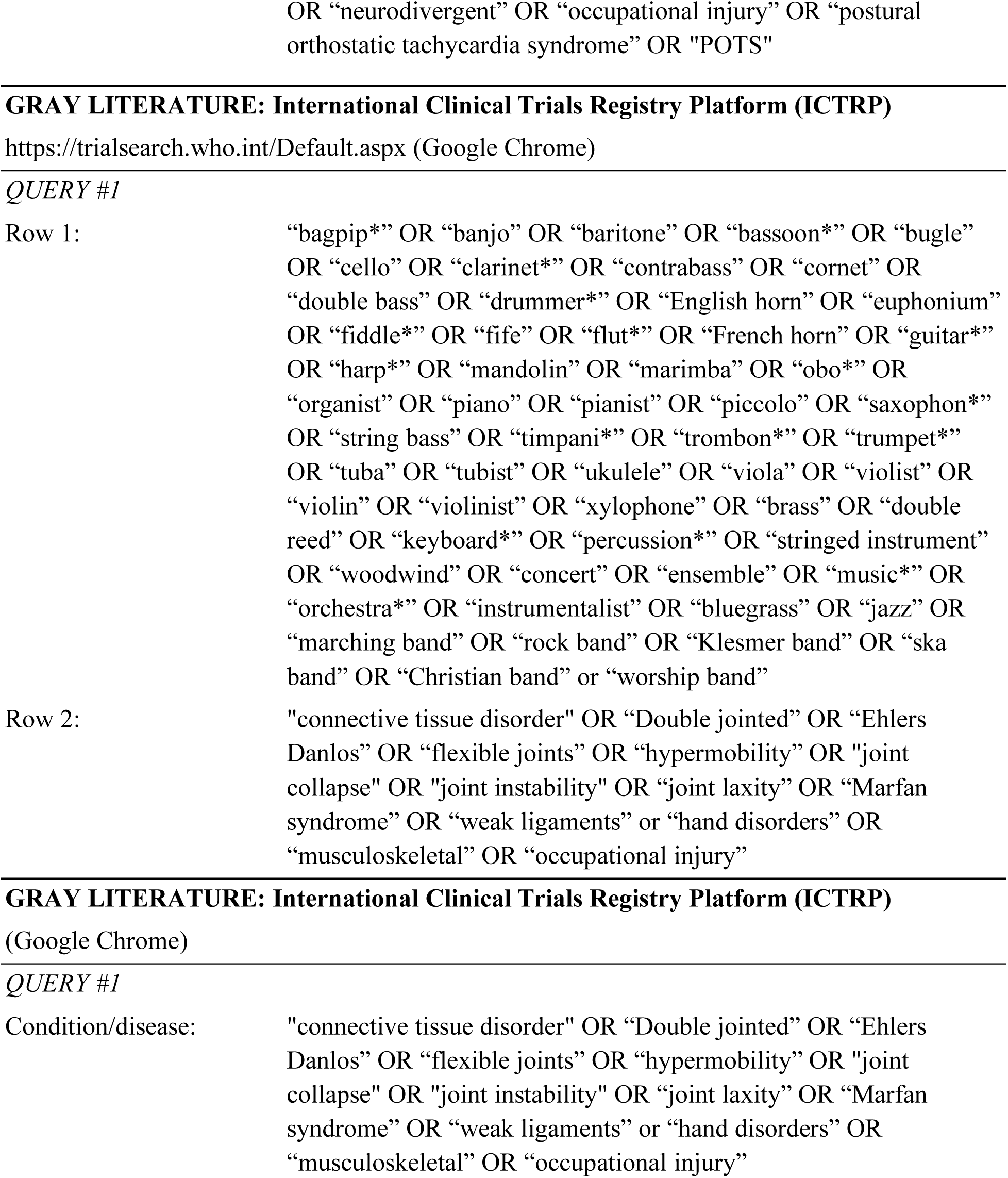
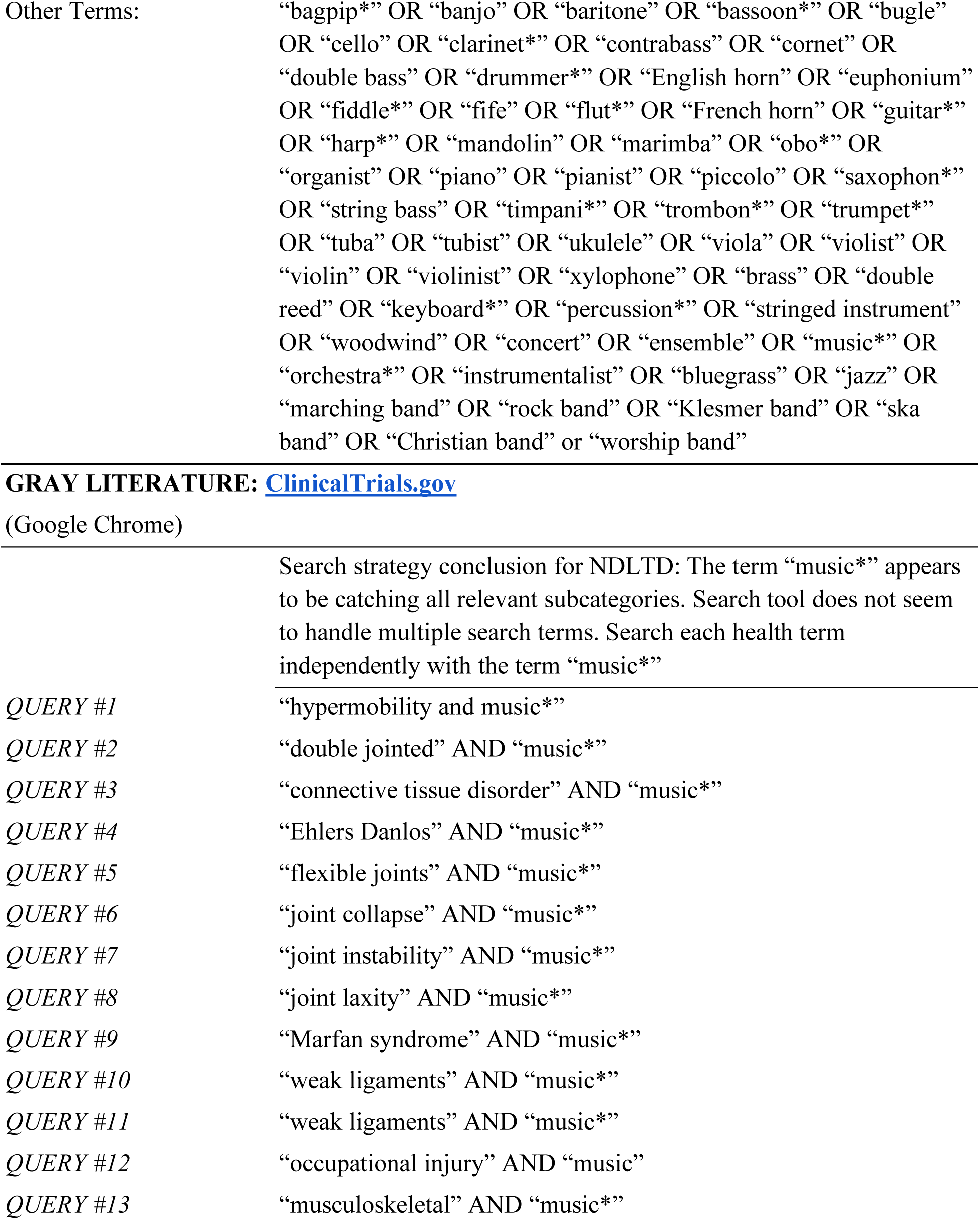
Search Query Strings.

### Appendix C

**Table C1.**
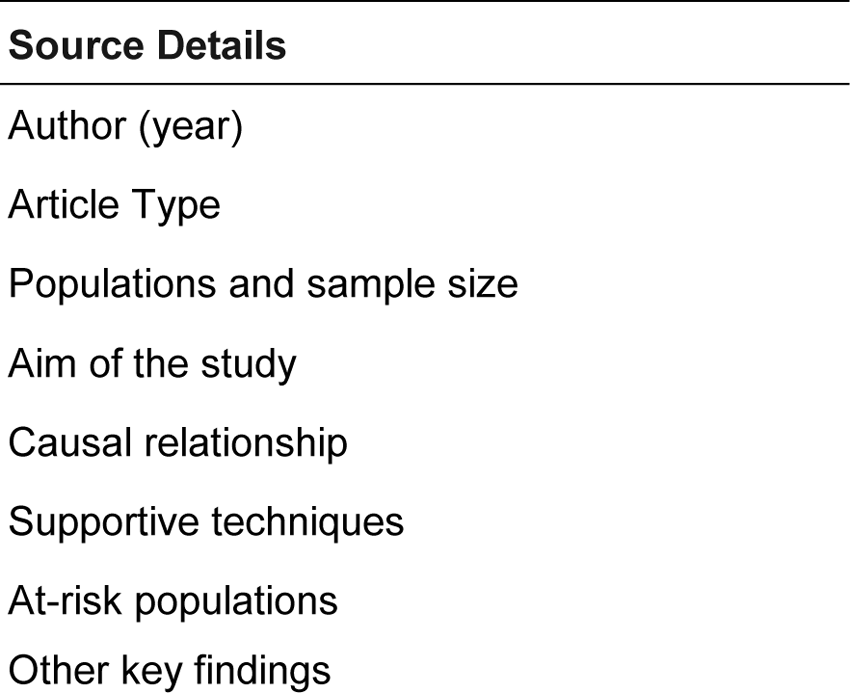
Initial Extraction Instrument.

**Table C2.**
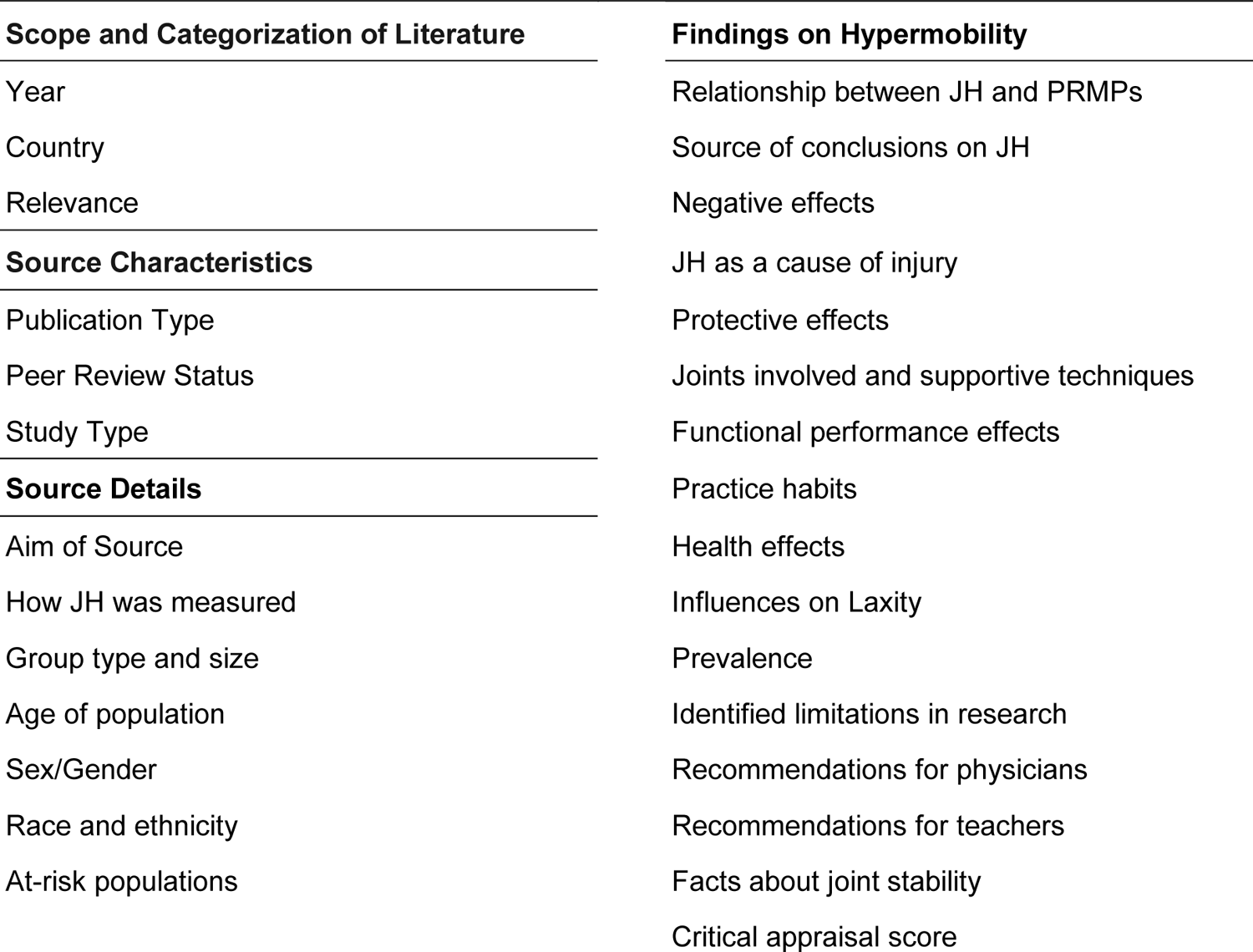
Adapted Extraction Instrument.

### Appendix D

**Table D1.**
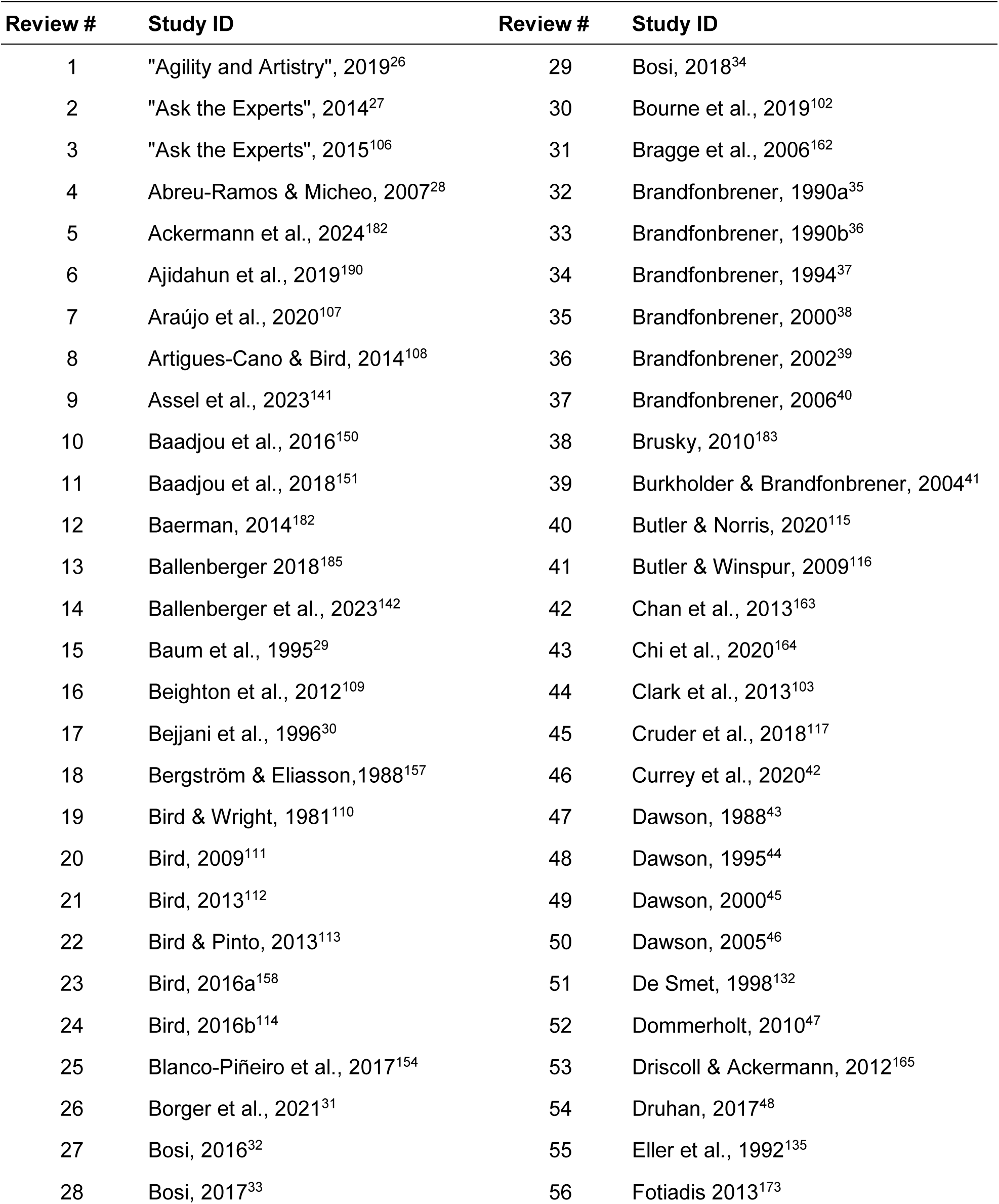
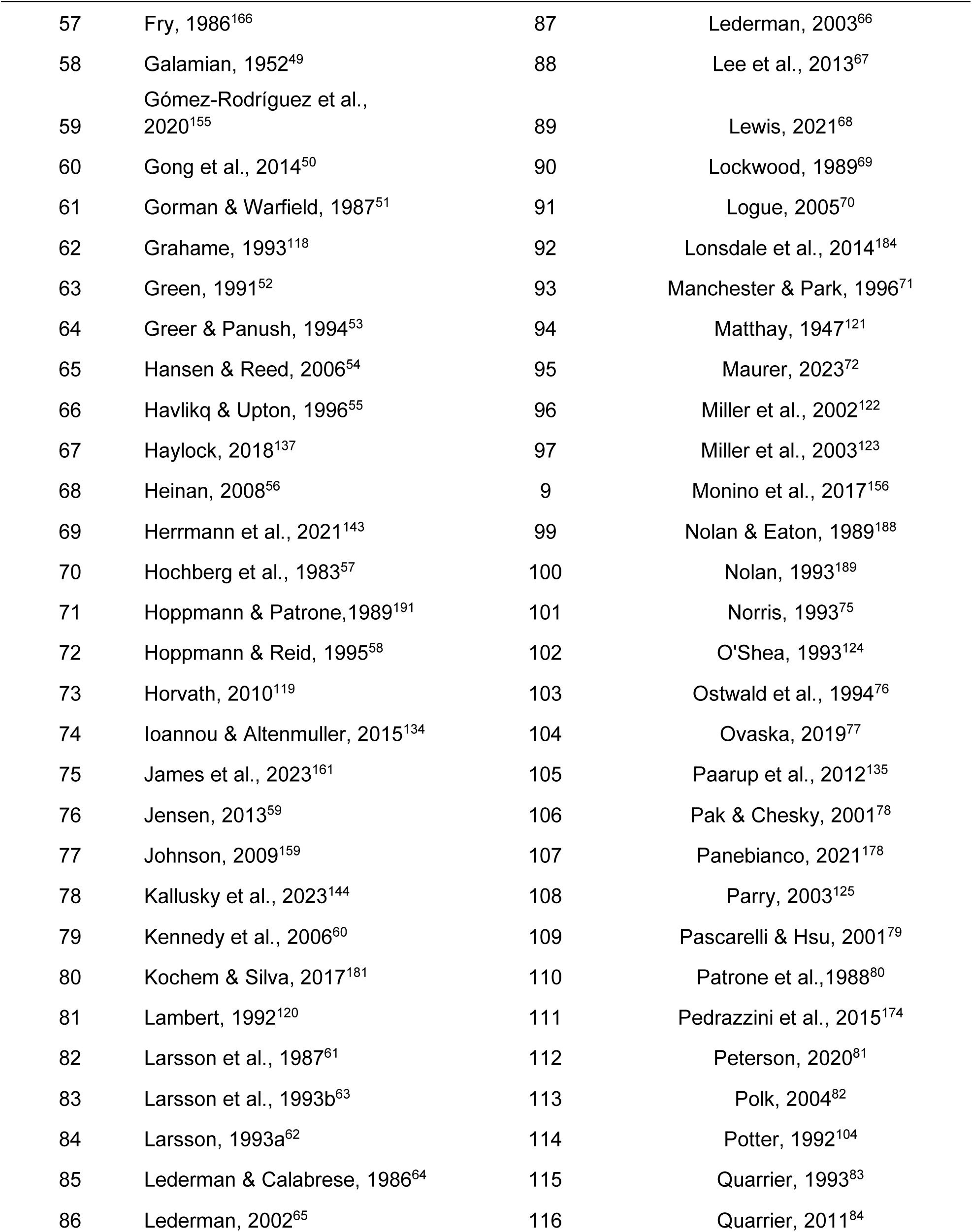
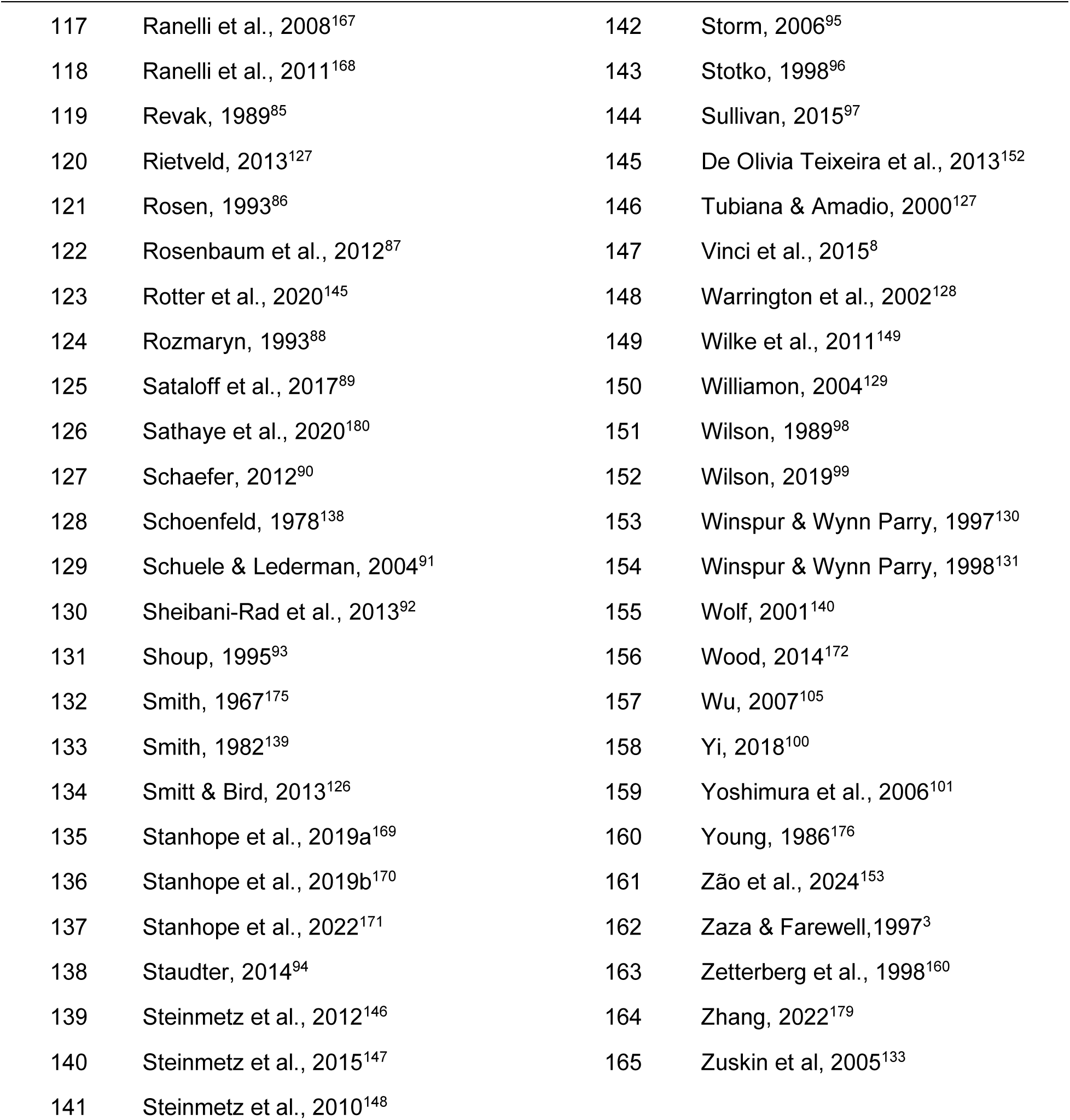
L1: All Included Sources (N = 165)

**Table D2.**
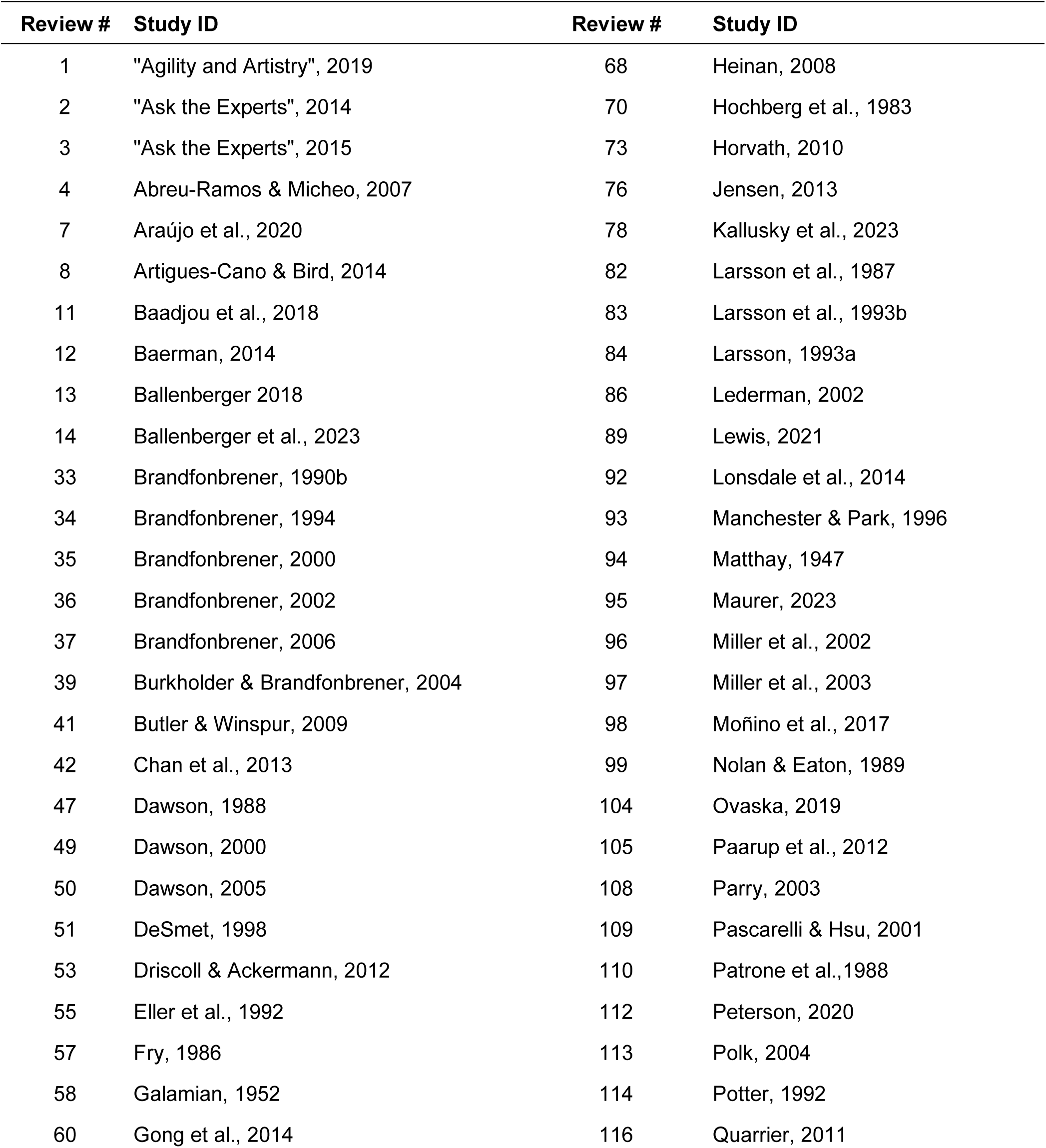
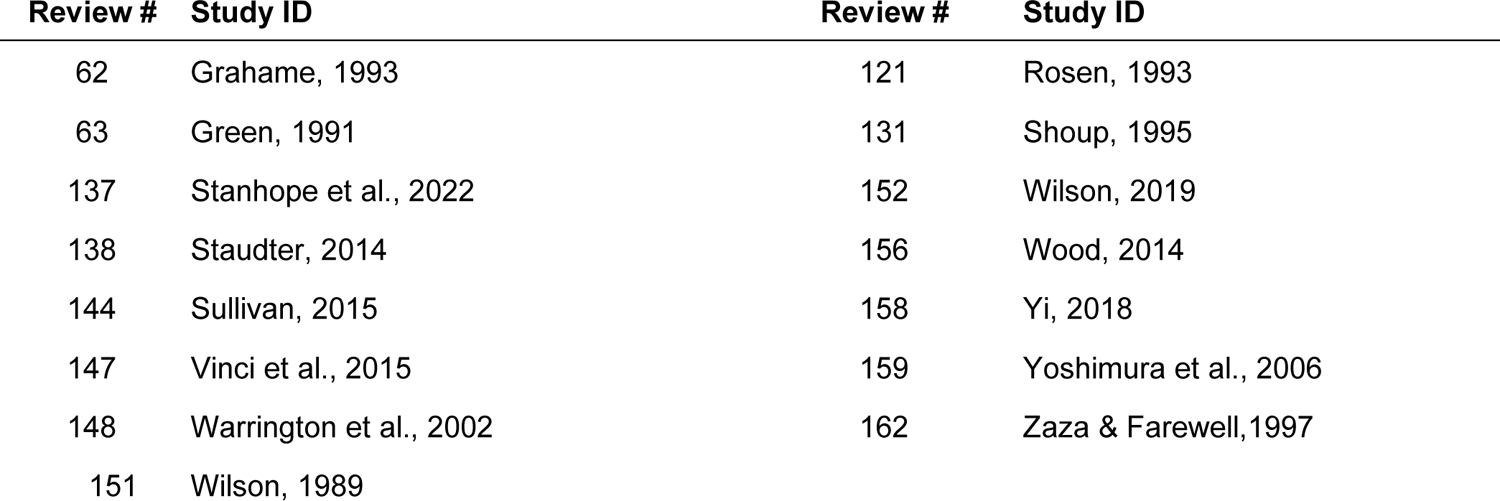
L2: Sources Containing Original Information on Hypermobility (n = 79)

**Table D3.**
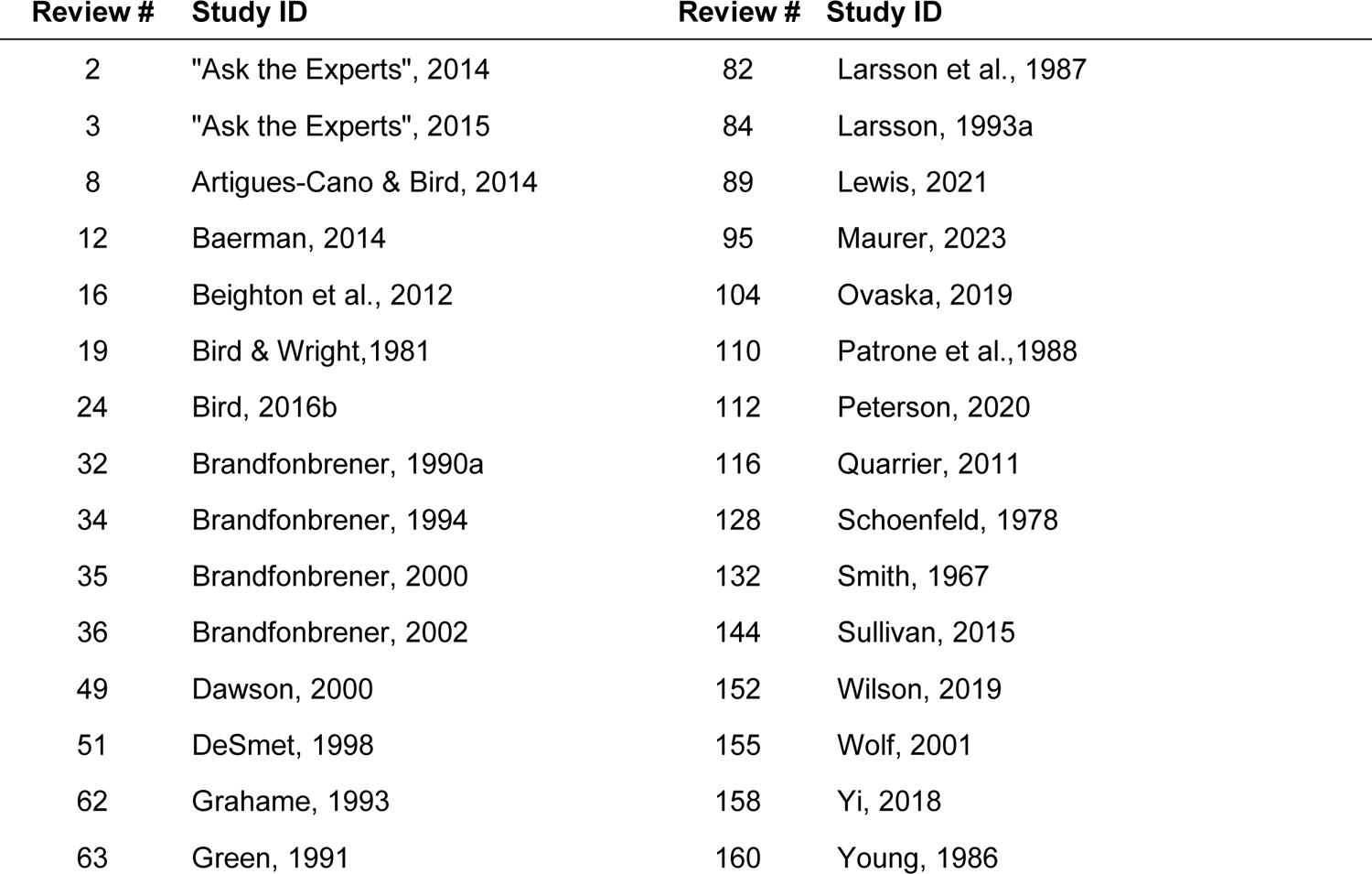
L3: Sources Focused on Hypermobility (n = 30)

#### Table D4: Original Prevalence Data

To avoid duplication of data specific to joint hypermobility prevalence L2 sources were grouped into three subcategories, shown in Table 2.4. Each article that includes original data relevant to the subcategory, regardless of the direction of the outcome, is marked in the corresponding column with an “x”.

- L2.1 sources contain original data comparing joint hypermobility prevalence among biological males to females
- L2.2 sources contain original data comparing injured musicians with and without joint hypermobility.
- L2.3 sources contain original data comparing joint hypermobility prevalence among musicians to the general population.

**Table D4.** L2 Prevalence Data Originality.

| **Table D4** |  |  |  |  |
| --- | --- | --- | --- | --- |
| *L2 Prevalence Data Originality* |  |  |  |  |
| **Review #** | **Study ID** | **L2.1:**<br>**Male/Female** | **L2.2 Injury Status** | **L2.3:**<br>**Musician/General**<br>**population** |
| 4 | Abreu-Ramos & Micheo, 2007 | x |  |  |
| 7 | Araújo et al., 2020 | x | x |  |
| 8 | Artigues-Cano & Bird, 2014 |  |  | x |
| 19 | Bird & Wright, 1981 | x | x | x |
| 32 | Brandfonbrener, 1990a | x | x | x |
| 33 | Brandfonbrener, 1990b |  | x | x |
| 35 | Brandfonbrener, 2000 | x | x |  |
| 36 | Brandfonbrener, 2002 | x | x | x |
| 39 | Burkholder & Brandfonbrener, 2004 | x | x | x |
| 42 | Chan et al., 2013 |  | x | x |
| 51 | DeSmet, 1998 | x | x |  |
| 53 | Driscoll & Ackermann, 2012 | x | x | x |
| 82 | Larsson et al., 1987 | x |  |  |
| 83 | Larsson et al., 1993b | x | x |  |
| 84 | Larsson, 1993a | x | x |  |
| 92 | Lonsdale et al., 2014 |  | x | x |
| 93 | Manchester & Park, 1996 |  | x | x |
| 96 | Miller et al., 2002 |  |  | x |
| 97 | Miller et al., 2003 |  |  | x |

| **Review #** | **Study ID** | **L2.1:**<br>**Male/Female** | **L2.2 Injury Status** | **L2.3:**<br>**Musician/General**<br>**population** |
| --- | --- | --- | --- | --- |
| 98 | Moñino et al., 2017 |  | x |  |
| 105 | Paarup et al., 2012 | x | x |  |
| 108 | Parry, 2003 |  | x | x |
| 109 | Pascarelli & Hsu, 2001 |  | x | x |
| 116 | Quarrier, 2011 | x |  |  |
| 131 | Shoup, 1995 |  | x | x |
| 147 | Vinci et al., 2015 |  | x | x |
| 148 | Warrington et al., 2002 | x |  |  |
| 159 | Yoshimura et al., 2006 |  | x | x |

### Appendix E

Other key findings

**Table E1.**
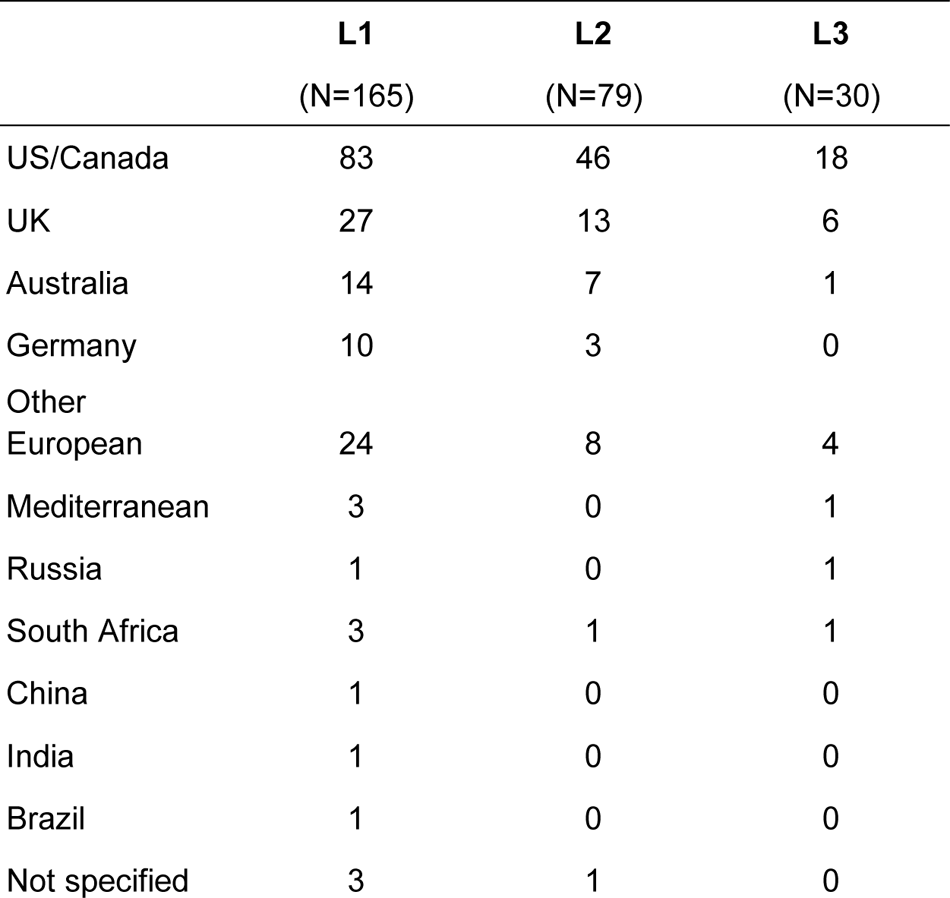
Country of Origin.

**Table E2.** Age Distribution.

|  | **L2** | **L3** |
| --- | --- | --- |
|  | (N=79) | (N=30) |
| Under 12 | 8 | 1 |
| 12-18 | 14 | 4 |
| 18-24 | 34 | 9 |
| 24-40 | 28 | 10 |
| 40-60 | 16 | 5 |
| 60+ | 12 | 4 |
| Not specified | 34 | 15 |

**Table E3.** Performance Setting.

|  | **L2** | **L3** |
| --- | --- | --- |
|  | (N=79) | (N=30) |
| Primary/Secondary | 10 | 2 |
| Higher Education | 28 | 10 |
| Amateur | 8 | 3 |
| Professional | 22 | 8 |
| Not Specified | 31 | 11 |

**Table E4.** Music Genre and Instrument Performed.

|  | **L2** | **L3** |
| --- | --- | --- |
|  | (N=79) | (N=30) |
| Classical | 19 | 8 |
| Jazz | 4 | 1 |
| Traditional | 1 | 0 |
| String | 9 | 10 |
| Keyboard | 6 | 1 |
| Flute | 4 | 2 |
| Woodwind | 4 | 3 |
| Guitar | 1 | 1 |
| Percussion | 1 | 1 |
| Bagpipe | 1 | 0 |
| Not Specified | 39 | 10 |

